# SAMSN1 causes sepsis immunosuppression by inducing macrophages to express coinhibitory molecules that causes T cell exhaustion via KEAP1-NRF2 signaling

**DOI:** 10.1101/2024.10.03.24314746

**Authors:** Yao Li, Tingting Li, Fei Xiao, Lijun Wang, Xuelian Liao, Yan Kang, Wei Zhang

## Abstract

Immunosuppression has been found to be closely related to the pathogenesis of sepsis, but the underlying mechanisms have not yet been fully elucidated. In this study, we identified that SH3 domain and nuclear localization signals 1 (SAMSN1), a gene encoding a putative adaptor protein, plays an important role in immunosuppression in sepsis. The expression of SAMSN1 was significantly increased in patients with sepsis and was positively correlated with sepsis mortality. When sepsis occurs, the number of monocyte-macrophages increases significantly, among which SAMSN1 is highly expressed. SAMSN1 binds to KEAP1, causing NRF2 to dissociate from the KEAP1-NRF2 complex and translocate into the nucleus, promoting the transcription of co-inhibitory molecules CD48/CD86/CEACAM1, which bind to their corresponding receptors 2B4/CTLA4/TIM3 on the surface of T cells, inducing T cell exhaustion. SAMSN1 blockade alleviated organ injuries and improved survival of septic mice. Our study reveals a novel mechanism that triggers immunosuppression in sepsis and may provide a candidate molecular target for sepsis immunotherapy.

## Introduction

Sepsis is currently defined as a life-threatening organ dysfunction caused by dysregulated host response to infection and is by far the most common cause of admission in intensive care units ^1–3^. Despite years of efforts, the mortality rate of sepsis remains high at approximately 22.5%. Rudd and colleagues analyzed multiple cause-of-death data from 109 million individual death records and a total of 48.9 million cases of sepsis were recorded worldwide, of which 11 million sepsis-related deaths were reported, representing 19.7% of all global deaths ^4^.

Excessive inflammatory response has long been considered to be at the core of the pathogenesis of sepsis. Unfortunately, all clinical trials targeting inflammatory mediators have failed over the past few decades, suggesting that inflammation alone does not fully reflect the host response during sepsis. The 2016 Third International Consensus Definition of Sepsis and Septic Shock (Sepsis-3) proposed that dysregulated host response caused by infection, rather than inflammation, is the core of the pathogenesis of sepsis, and immunosuppression is an important component of this dysregulated response^5–7^. Various manifestations of immunosuppression in sepsis include increased secretion of immunosuppressive factors (IL-4, IL-10, etc.), increased expression of immunosuppressive surface molecules (PD-L1, CTLA-4, etc.), increased immunosuppressive cells (mainly Treg cells and myeloid-derived suppressor cells), and decreased adaptive immune cells (mainly B cells and T cells)^5^. However, although immunosuppression in sepsis has been widely studied, it is still unclear how it is triggered, what changes it causes in the host’s immune response, and what the molecular mechanisms behind it are. When sepsis occurs, there may be some specific mechanisms that trigger this immunosuppression. If the key genes involved in these mechanisms can be identified, it will fill in the gaps in our understanding of the pathogenesis of sepsis and provide innovative diagnostic markers and targets for sepsis intervention.

In this study, we used the following strategy to identify potential genes that may affect the pathogenesis of sepsis: first, we analyzed the public gene sequencing database to screen for genes with significant expression changes during sepsis; second, we used prospective RNA-Seq data for external validation; third, we performed correlation analysis between the screened candidate gene and the clinical indicators, severity and survival rate of sepsis patients. We identified SH3 domain and nuclear localization signals 1 (SAMSN1), a gene encoding a putative adaptor protein, whose expression is significantly increased during sepsis. Higher SAMSN1 levels are positively correlated with sepsis severity and mortality. Through gene editing, flow cytometry and functional experiments, we found that SAMSN1 was mainly expressed in monocytes-macrophages and promoted the expression of co-inhibitory molecules CD48, CD86 and CEACAM1, which bind to their corresponding receptors 2B4, CTLA4, TIM3 on the surface of T cells, inducing T cell exhaustion. SAMSN1 binds to Kelch-like ECH-associated protein 1 (KEAP1), causing the transcription factor Nuclear factor erythroid 2-related factor 2 (NRF2) to dissociate from the KEAP1-NRF2 complex, NRF2 then translocates into the nucleus and initiates the transcription of downstream target genes including CD48, CD86, and CEACAM1. Our study reveals a novel mechanism that triggers immunosuppression in sepsis, and may provide translational opportunities for the design of sepsis immunotherapy.

## Results

### Bioinformatics analysis and external validation revealed that SAMSN1 expression was increased in patients with sepsis and was negatively correlated with survival

To comprehensively screen the gene expression of immune responses when sepsis occurs, we first used GEO database to analyze the differences in gene expression between sepsis patients and healthy volunteers. By constructing search terms, we retrieved a total of 56 public sequencing data sets related to sepsis between 2016 and 2021, and selected 3 datasets that met the requirements for analysis (Supplementary Table 1, 2). Setting the definition of differential genes to adjP value < 0.05 and |log Foldchange|>2, there were 209, 769, and 232 differentially expressed genes were screened from datasets GSE131761, GSE154918, and GSE139913, respectively, among which 32 genes were shared by these three datasets (Fig. 1a). Among the 32 genes, a gene named SH3 domain and nuclear localization signals 1 (SAMSN1) showed significantly increased expression in the sepsis patients in all three datasets (Fig. 1a, b).

**Figure 1.**
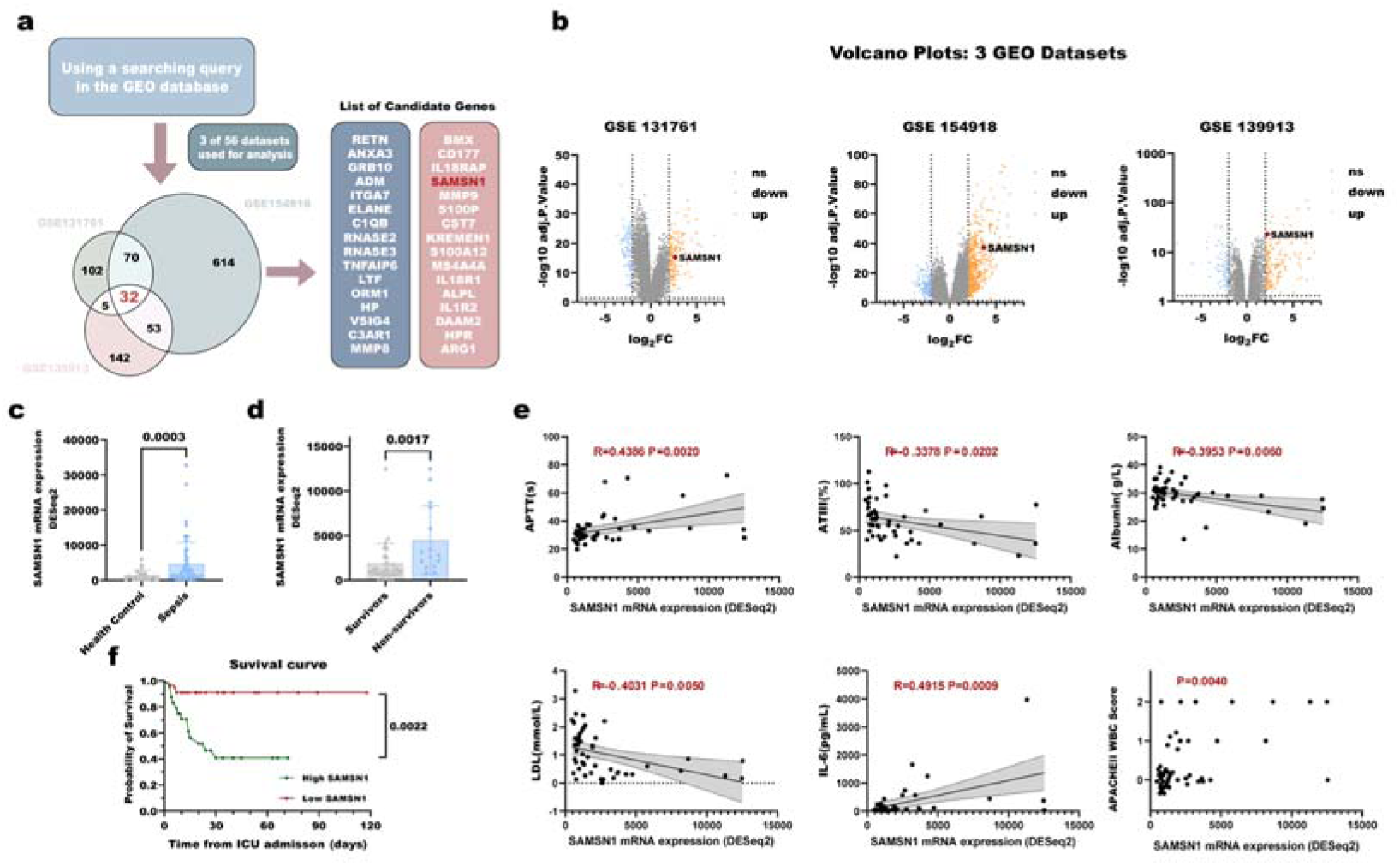
SAMSN1 expression was increased in patients with sepsis and was positively correlated with sepsis severity and mortality. **(a)** Flow chart of screening candidate genes related to sepsis using the public GEO database. **(b)** Volcano plots of DEGs in 3 GEO Datasets. Colored dots represent DEGs based on the criteria of adjusted p-value <L0.05 and |log2FC| >L2; orange: up-regulation, blue: downregulation, grey: non-significant mRNAs. Red dots specifically indicate SAMSN1. **(c)** RNA-Seq data from PBMCs from 59 sepsis patients and 27 healthy volunteers were used to externally validate the expression level of SAMSN1. **(d)** RNA-Seq results showed that the SAMSN1 levels of non-survivors (n=15) were significantly higher than those of survivors (n=32). The significance in **(c, d)** was assessed by *Mann-Whitney u test* for the data did not conform to a normal distribution. **(e)** Correlation analysis between SAMSN1 and clinical indicators APTT, ATIII, albumin, LDL and IL-6. SAMSN1 and APACHEII score were analyzed using Kruskal-Walli test. **(f)** Kaplan-Meier survival analysis was used to analyze the correlation between SAMSN1 expression and survival in patients with sepsis. Sepsis patients were divided into SAMSN1 high expression group (Read account > 1352, n=24) and low expression group (Read account ≤ 1352, n=23). The significance in was assessed by *log-rank test*.

We used RNA-Seq data from peripheral blood mononuclear cells (PBMCs) from 59 sepsis patients and 27 healthy volunteers for external validation. The expression change trend of SAMSN1 was consistent with the trend derived from the analysis of public data sets, and the SAMSN1 mRNA level in sepsis patients was significantly higher than that in healthy controls (Fig. 1c). In addition, a markedly higher SAMSN1 level was observed in survivors than in non-survivors (Fig. 1d). There was a linear correlation between SAMSN1 and coagulation markers (APTT, ATIII), inflammation markers (IL-6, APACHEII WBC score), and metabolic markers (albumin, LDL) (Fig. 1e and Supplementary Table 3). Kaplan-Meier survival analysis showed that patients with higher SAMSN1 levels had a significantly higher mortality rates (Fig. 1f).

### Blockade of SAMSN1 improved the survival rate of septic mice by alleviating acute organ injuries

We used a classic mouse model, cecal ligation and puncture (CLP)-induced sepsis, to investigate the potential role of SAMSN1 in sepsis. We found that the mRNA level of SAMSN1 not only in PBMCs, but also in bone marrow, spleen, peripheral blood, and liver in the CLP group was significantly higher than that in the control group (Fig. 2a). We employed the CRISPR/Cas9 system to generate SAMSN1 knockout (*Samsn1^−/−^*) mice, which were ostensibly viable and fertile (Supplementary Fig. 1, 2 and Supplementary Materials 1). Weight-matched wild-type mice and *Samsn1^−/−^* mice underwent CLP surgery, and 14-day survival rates were recorded. The survival experiment showed that *Samsn1^−/−^* mice had an increased survival rate, fewer weight change, and a milder symptom score (Fig. 2b-d).

**Figure 2.**
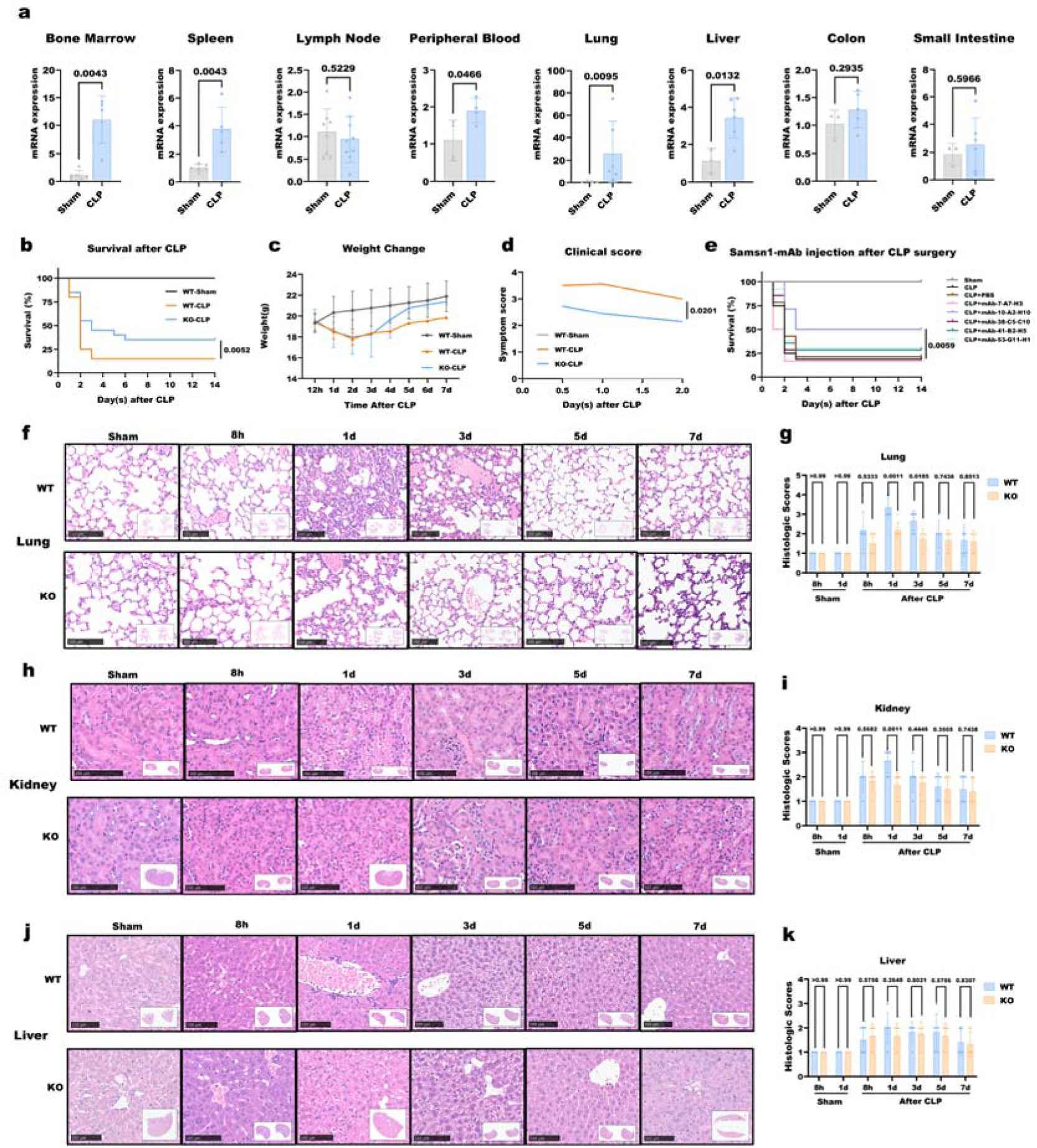
Blockade of SAMSN1 improved the survival rate of septic mice by alleviating acute organ injuries. **(a)** Comparison of SAMSN1 mRNA levels in different tissues and organs between control and septic mice by Realtime-RT-PCR. **(b)** Kaplan–Meier analysis of the survival rate of WT C57BL/6 mice and SAMSN1-KO mice after CLP surgery. **(c)** Body weight changes in WT and KO mice after CLP surgery. **(d)** Clinical scores after CLP surgery in WT and KO mice. In **(b-d)**, WT-Sham group, n=5; WT-CLP group, n=20; KO-CLP group, n=20. **(e)** Kaplan–Meier analysis of the therapeutic effects of 5 anti-SAMSN1 monoclonal antibodies on CLP mice. Sham, n=5; CLP group and CLP + mAbs groups, n=12-14. **(f, h, k)** H&E staining of lungs, kidneys, and livers at 8h, 1d, 3d, 5d, and 7d after CLP. **(g, i, k)** Injury scores of the lung, kidney, and liver of mice at 8h, 1d, 3d, 5d, and 7d after CLP surgery. Significance in **(a, d, g, i, k)** was assessed by *2-tailed Welch’s t test,* and significance in **(b, e)** was assessed by *Log-rank test*.

To verify the potential impact of intervening SAMSN1 on sepsis survival, we prepared 6 anti-SAMSN1 monoclonal antibody candidate strains, among which mAb-10-A2-H10 showed significant survival benefit (Fig. 2e and Supplementary Fig. 3). The lungs, kidneys, and livers were collected at 8h, 1d, 3d, 5d, and 7d after CLP surgery to assess acute organ injuries in sepsis. Histopathological scoring of H&E stained sections (Fig. 2f, h, j) ^8^ showed significant lung and kidney injuries after CLP, whereas no significant changes were found in the liver (Fig. 2g, i, k). The lungs of wild-type mice showed large hemorrhagic lithiasis and concomitant severe inflammatory infiltrates, whereas the corresponding *Samsn1^−/−^* mice had less severe damage (Fig. 2f, g). The wild-type mice showed obvious acute kidney injury (AKI) 1 day after CLP, characterized by nuclear vacuolar degeneration of renal tubular epithelial cells, necrotic detachment, and formation of intratubular casts (Fig. 2h). *Samsn1^−/−^* mice showed an alleviated renal injury, and this effect was most significant 1 day after CLP (Fig. 2i). ELISA showed that the inflammatory factors IL-6 and TNF-α gradually increased in the serum of mice after CLP, while IL-10 showed an opposite trend, but no significant difference was found between wild-type and *Samsn1^−/−^* mice (Supplementary Fig. 4).

### SAMSN1 blockade enhances macrophage proliferation, phagocytosis, and clearance of bacteria from the blood

SAMSN1 was first identified in leukemia cells and mast cells. We analyzed the expression of SAMSN1 in PBMCs of sepsis patients in the dataset GSE151263 and found that SAMSN1 was expressed on various types of immune cells, but mainly in monocytes-macrophages (Supplementary Fig. 5). We therefore chose murine mono-macrophage cell line RAW264.7 as a model cell and generated SAMSN1-KO RAW264.7 cells using Crisper-cas9 system to investigate the role of SAMSN1 (Fig. 3a and Supplementary Fig. 6-8).

**Figure 3.**
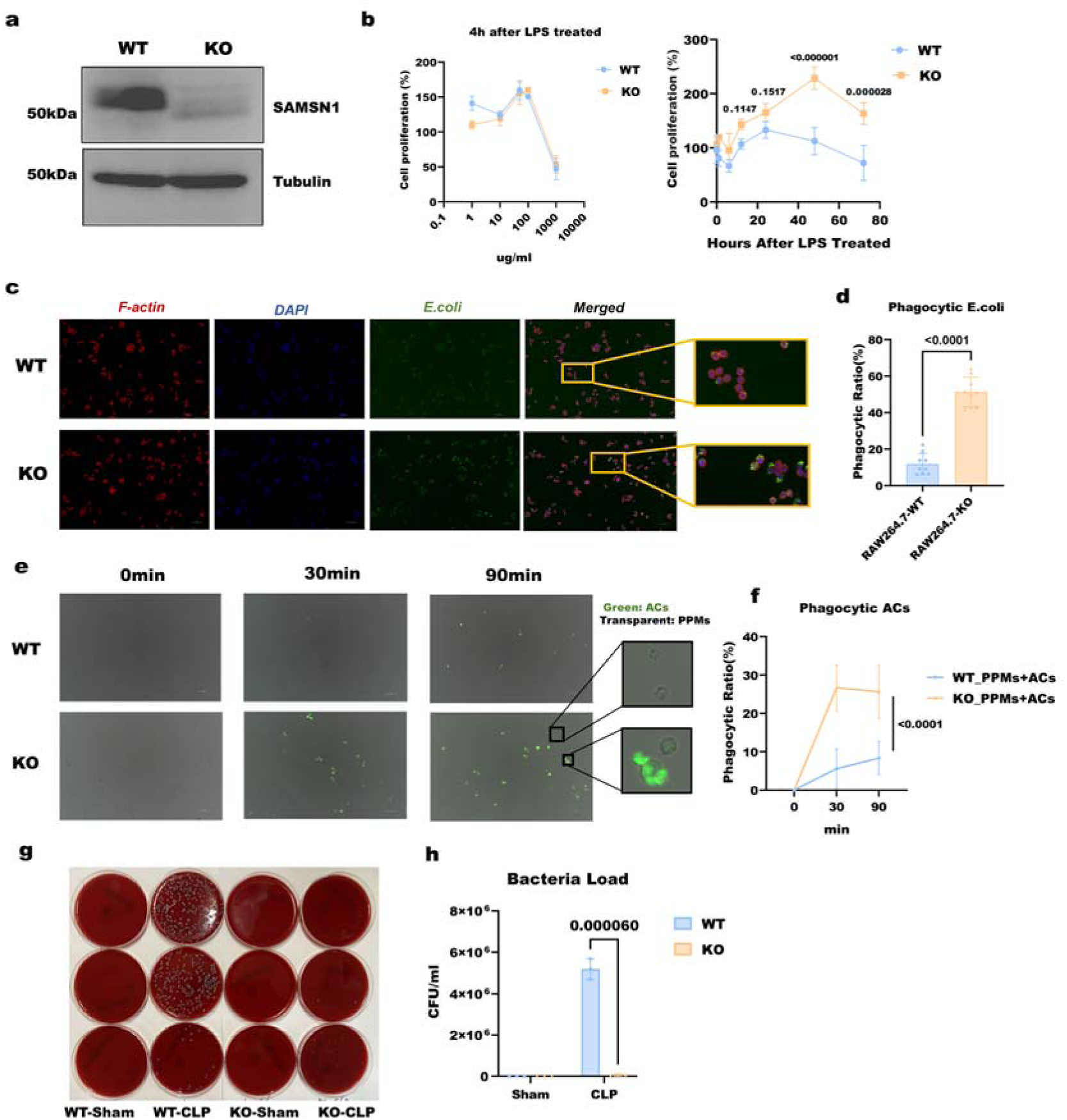
SAMSN1 knockout enhances macrophage proliferation, phagocytosis, and clearance of bacteria from the blood. **(a)** Western blot (WB) was used to detect whether SAMSN1 was successfully knocked out in RAW264.7 cells. WT: wild type RAW264.7. KO: SAMSN1-KO RAW264.7. **(b)** Left panel: the proliferation of WT and SAMSN1-KO cells was detected after 4 hours of LPS treatment with different concentrations (0, 1, 10, 50, 100, 1000ug/ml). Right panel: after adding 50ug/ml LPS, the proliferation of WT and SAMSN1-KO cells was detected at 0, 1, 6, 12, 24, 48, and 72 hours (n=3). **(c)** RAW264.7 cells phagocytose E. coli. Red: F-actin; Blue: DAPI; Green: E. coli. RAW264.7 cells were co-cultured with E. coli at a ratio of 1:10 for 60 minutes. **(d)** Quantification of phagocytotic ratio % (n=9). **(e)** Analysis of phagocytosis of apoptotic Jurkat cells by primary peritoneal macrophages. The prepared apoptotic Jurkat cells (green) (ACs) and primary peritoneal macrophages (transparent) (PPMs) were co cultured in 1:1 ratio for 0,30 and 90 minutes. The enlarged images show PPMs with or without phagocytosis of ACs. **(f)** Quantification of the ratio of macrophages phagocytosing apoptotic cells % (n=9). **(g)** Blood agar plate assay to measure bacterial load in peripheral blood of WT and KO mice after CLP surgery. **(g)** Quantification of bacterial load in each group (n=3). Significance was assessed by *2-tailed Welch’s t test*.

We first tested the stimulating effect of different doses of lipopolysaccharide (LPS) on macrophages in vitro. When the LPS dose reached 50 μg/ml, the proliferation effect on RAW264.7 cells was the strongest, while higher doses were toxic to cells (Fig. 3b). Under the stimulation of 50 μg/ml LPS, the proliferation of SAMSN1-KO cells was significantly higher than that of WT cells, suggesting that SAMSN1 may negatively regulate the proliferation of macrophages (Fig. 3b).

Phagocytosis is the essential function of macrophages to exert their biological effects. We used GFP-labeled *E. coli* as a model bacterium for phagocytosis experiments and found that the phagocytic ability of SAMSN1-KO cells was significantly stronger than that of WT cells (Fig. 3c, d). We also isolated primary peritoneal macrophages from WT and *Samsn1^-/-^*mice and tested their ability to phagocytose pyroptotic cells. The primary macrophages were co-cultured with apoptotic Jurkat cells, an immortalized T lymphocyte line. Fluorescence co-localization experiments showed that macrophages derived from *Samsn1^-/-^* mice had a stronger ability to phagocytose apoptotic cells (Fig. 3e, f and Supplementary Fig. 9). Blocking SAMSN1 function promotes macrophage proliferation and phagocytosis, suggesting that it may help clear invading bacteria in sepsis. We collected blood samples from septic mice 12 hours after CLP surgery for blood culture testing. Compared with WT mice that underwent CLP surgery, bacteria in the blood of *Samsn1^-/-^*mice were largely eliminated after CLP (Fig. 3g, h).

### SAMSN1 affects immune cell populations in normal and septic mice

To evaluate the effect of SAMSN1 on immune cell populations in septic mice, we collected peripheral blood, spleen, and bone marrow samples 12 h after CLP for flow cytometric analysis. The number and proportion of CD3^+^, CD4^+^ and CD8^+^ T cells in the spleen and bone marrow of *Samsn1^-/-^*mice were higher than that of wild-type mice (Fig. 4a-f), but myeloid cells in *Samsn1^-/-^* mice were lower than that of WT mice (Supplementary Fig. 10). For B cells, the number and proportion of B220^+^ and B220^+^CD5^+^ B cells increased slightly in the peripheral blood and spleen of *Samsn1^-/-^*mice, but did not significantly change in the bone marrow (Supplementary Fig. 10). These results indicate that in *Samsn1^-/-^*mice, the number and proportion of T cells significantly increased, B cells increased slightly, while myeloid cells decreased.

**Figure 4.**
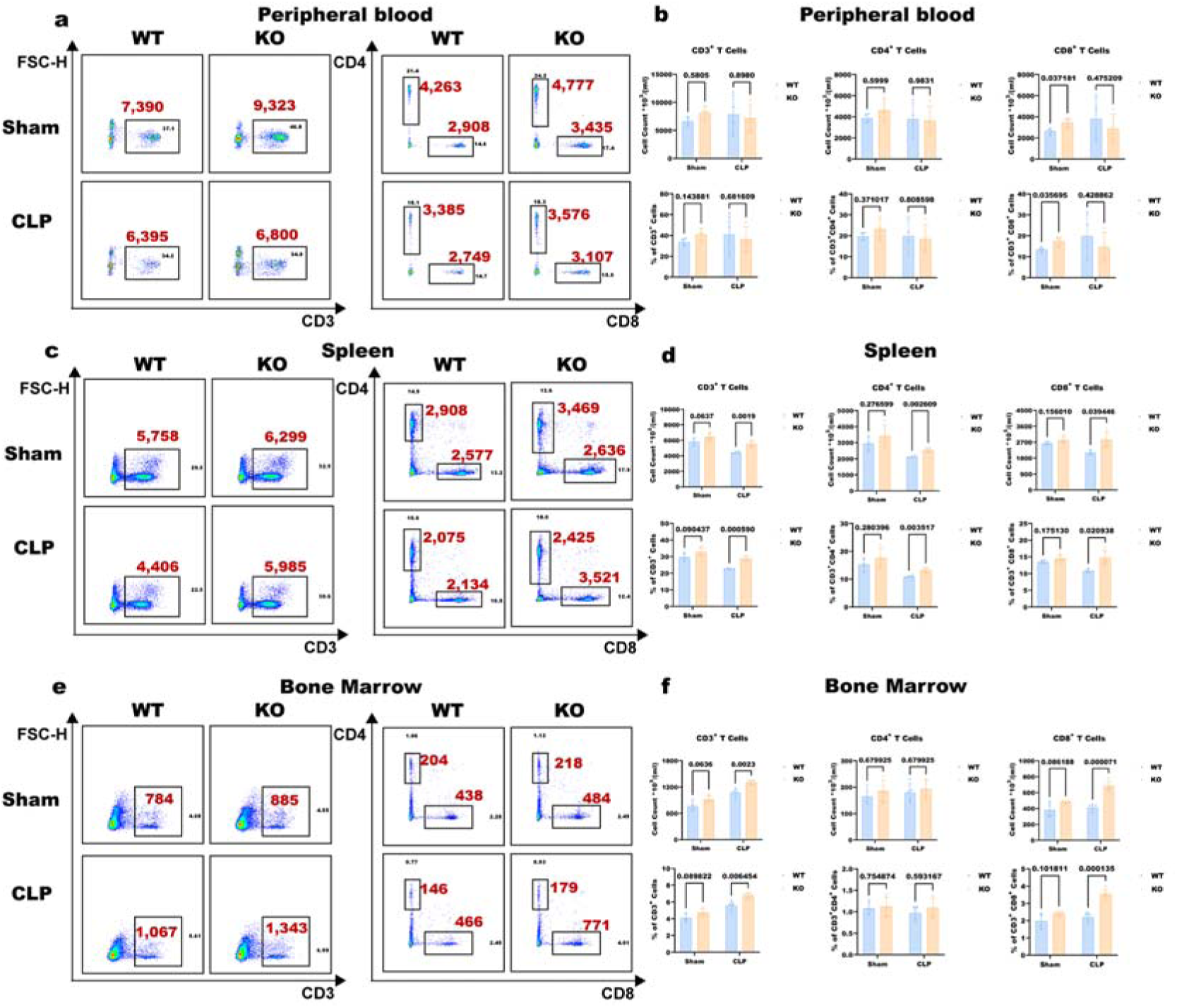
SAMSN1 affects T cell populations in normal and septic mice. **(a, c, e)** Flow cytometric analysis of T cell populations (CD3^+^, CD3^+^CD4^+^, CD3^+^CD8^+^ T cells) in peripheral blood, spleen, and bone marrow of WT and SAMSN1-KO mice after sham or CLP surgery. Samples were taken 12 hours after Sham or CLP surgery. Red text highlights the cell number in each group. **(b, d, f)** Quantification of changes in T cell proportions and cell numbers. The comparison between two adjacent groups was performed using *2-tailed Welch’s t test* (n=3).

### SAMSN1 mediates the inhibition of T cells by macrophages through direct contact

Considering that SAMSN1 gene deletion increased the number of T cells and enhanced the ability of CD11b^+^ myeloid cells to clear bacteria, we speculated that SAMSN1 might mediate the crosstalk between myeloid cells and T cells. To verify this, we co-cultured RAW264.7 cells with primary T cells, and performed flow cytometry to detect changes in the cells after 12 hours of co-culture. We used two co-culture modes. First, the two cells were directly cultured in the same culture dish, so that macrophages could directly contact T cells; second, cells were cultured in the upper and lower chambers of the Transwell system to detect whether secreted factors produced by macrophages affect T cells. Flow cytometry showed that the number of T cells after direct co-culture with SAMSN1-KO RAW264.7 cells for 12 hours was significantly higher than that after co-culture with WT cells (Fig. 5a-e). However, RAW264.7 cells cultured in the upper chamber of the Transwell did not affect the number of T cells in the lower chamber (Fig. 5f-j). This suggests that macrophages are likely to reduce the number of T cells through direct contact, and knockout of the SAMSN1 gene inhibits this effect, resulting in a significant increase in CD3^+^CD4^+^ and CD3^+^CD8^+^ T cells.

**Figure 5.**
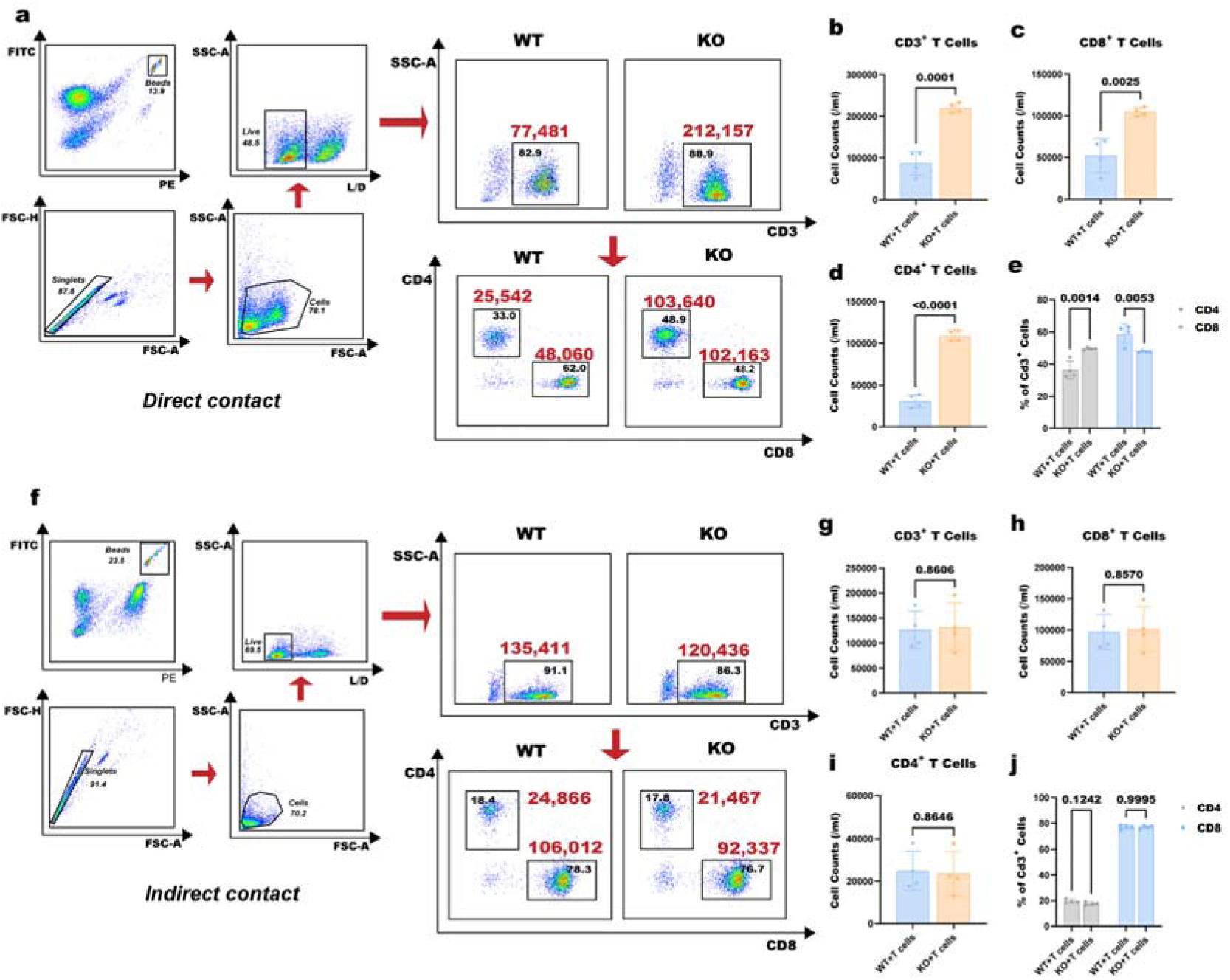
SAMSN1 mediates the inhibition of T cells by macrophages through direct contact. **(a)** Flow cytometry analysis after 12 h of direct WT or SAMSN1-KO RAW264.7 cells and T cell co-culture. Red text highlights the cell number in each group. **(b-e)** Quantify the changes in the number of CD3^+^, CD4^+^, and CD8^+^ T cells. **(f)** WT or SAMSN1-KO RAW264.7 cells were cultured in the upper chamber of the Transwell system, and T cells were cultured in the lower chamber. Flow cytometry was used to detect changes in the T cell population. **(g-j)** Quantify the changes in the number of CD3^+^, CD4^+^, and CD8^+^ T cells. Significance in **(b-d)** and **(g-i)** was assessed by *2-tailed Welch’s t test*. Statistical analysis of **(e, j)** was performed using *2-way ANOVA test* (n=4).

### The SAMSN1-mediated T cell exhaustion may be through upregulating the expression of CD48, CD86, and CEACAM1 on macrophages and binding to the corresponding receptors on T cells

To explore the effect of SAMSN1 in macrophages, we compared the expression differences between SAMSN1-KO RAW264.7 cells and WT cells using RNA-Seq. Principal component analysis (PCA) showed that the expression characteristics of SAMSN1-KO RAW264.7 and WT cells were clearly divided into 2 groups (Fig. 6a). There was a total of 518 differentially expressed genes, of which 319 gene expressions were upregulated and 199 genes downregulated in SAMSN1-KO cell (Fig. 6b). Gene Ontology (GO) analysis showed that the genes identified from KO vs. WT cells were most significantly enriched in the terms “defense response to other organism”, “lymphocyte activation”, and “T cell activation” (Fig. 6c). A total of 134 genes related to immune response were screened out from 518 differential genes, including 42 down-regulated genes and 92 up-regulated genes (Fig. 6d, Supplementary Fig. 11). Three genes that may mediate the interaction between macrophages and T cells were screened out, namely CD48, CD86 and CEACAM1, which are usually expressed on antigen-presenting cells (APCs) and can bind to inhibitory receptors 2B4 ^9^,CD152 ^10^ and TIM3 ^11^ on the surface of T cells, respectively. The combination of the three pairs of molecules transmits negative immune signals and mediates T cell exhaustion ^12^. We also analyzed the expression of other T cell inhibitory receptors and ligands, but these genes showed no differences between groups (Supplementary Fig. 12). Flow cytometry was used to verify the RNA-Seq results, showing that the expression of CD48, CD86 and CEACAM1 on the surface of SAMSN1-KO RAW264.7 cells was significantly reduced compared with that of WT cells (Fig. 6f-g). These results suggest that SAMSN1 may mediate T cell exhaustion by upregulating the expression of CD48, CD86 and CEACAM1 on macrophages and binding to their corresponding receptors on the surface of T cells.

**Figure 6.**
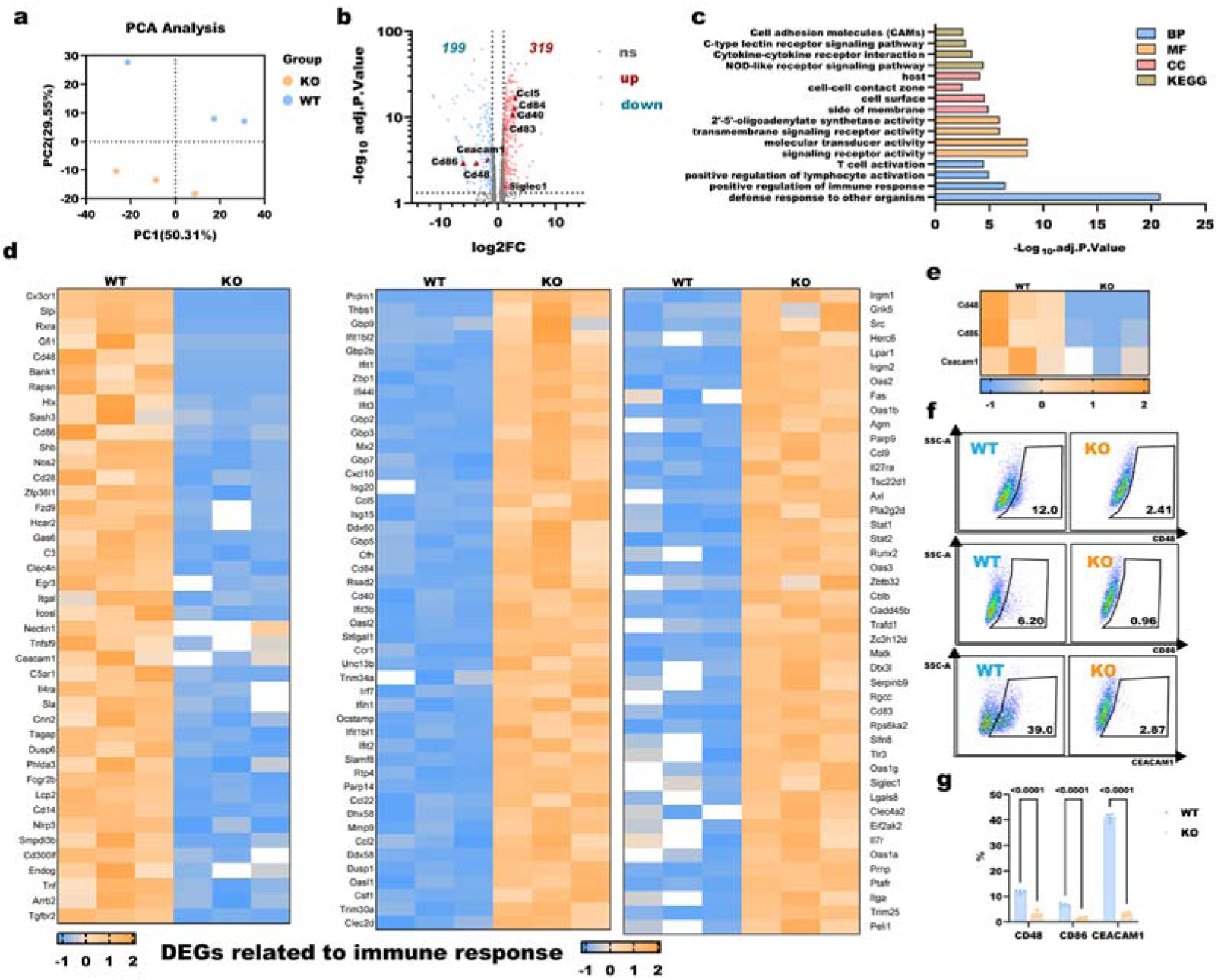
The SAMSN1-mediated T cell exhaustion may be through upregulating the expression of CD48, CD86, and CEACAM1 on macrophages and binding to the corresponding receptors on T cells.. **(a)** Principal component analysis (PCA) of RNA-Seq results of WT (blue) and SAMSN1-KO (orange) RAW264.7 cells. **(b)** Volcano plot analysis of gene expression differences. Adjusted p-value <L0.05 and |log2FC| >L1 were considered as genes with significant differences. **(c)** The Gene Ontology (GO: including BP, MF, CC) and Kyoto Encyclopedia of Genes and Genomes (KEGG) enrichment analysis. **(d)** Heatmap analysis showed significantly down-regulated genes (42) and up-regulated genes (92) after SAMSN1 knockout in RAW264.7 cells. **(e)** Heatmap showed that three immune co-inhibitory molecules CD48, CD86 and CEACAM1 were significantly decreased in SAMSN1-KO cells. **(f, g)** Flow cytometry showed that the expression of CD48, CD86, and CEACAM1 on the surface of SAMSN1-KO RAW264.7 cells was significantly lower than that of WT cells. Statistical analysis in **(g)** was performed using *2-way ANOVA test* (n=4).

### *Samsn1^-/-^* mice have decreased T cell exhaustion-related receptors expression and enhanced T cell activation and signaling

We next examined the corresponding inhibitory receptors on T cells. The expression of 2B4, CD152 and TIM3 on the surface of CD3^+^ T cells in the spleen of *Samsn1^-/-^*mice was markedly reduced, but no significant changes were found in the peripheral blood and bone marrow. (Fig. 7a-b). Similar to this expression trend, after CLP-surgery, 2B4, CD152, and TIM3 on T cells in the spleen were reduced in *Samsn1^-/-^* mice, while there were no obvious changes in peripheral blood and bone marrow (Supplementary Fig. 13). We then examined the expression of activating receptors CD69 and CD25 on splenic T cells. Under normal conditions, there was no significant difference in the expression of CD69 in splenic T cells of WT and *Samsn1^-/-^*mice. However, under sepsis, the expression of CD69 in splenic T cells of *Samsn1^-/-^*mice was significantly increased (Fig. 7c-d). As for the expression of CD25, no significant change was observed after CLP surgery or between WT and *Samsn1^-/-^* mice (Fig. 7e-f). ZAP70 and LCK are two non-receptor tyrosine kinases, playing a crucial role in the regulation of TCR signaling critical for T cell functions. Western blot (WB) detection showed that the phosphorylation level of ZAP70 and LCK in splenetic T cells of *Samsn1^-/-^* mice significantly increased in both normal and septic states (Fig. 7g-h, Supplementary Fig. 14), suggesting that the activity of T cells in *Samsn1^-/-^* mice is increased.

**Figure 7.**
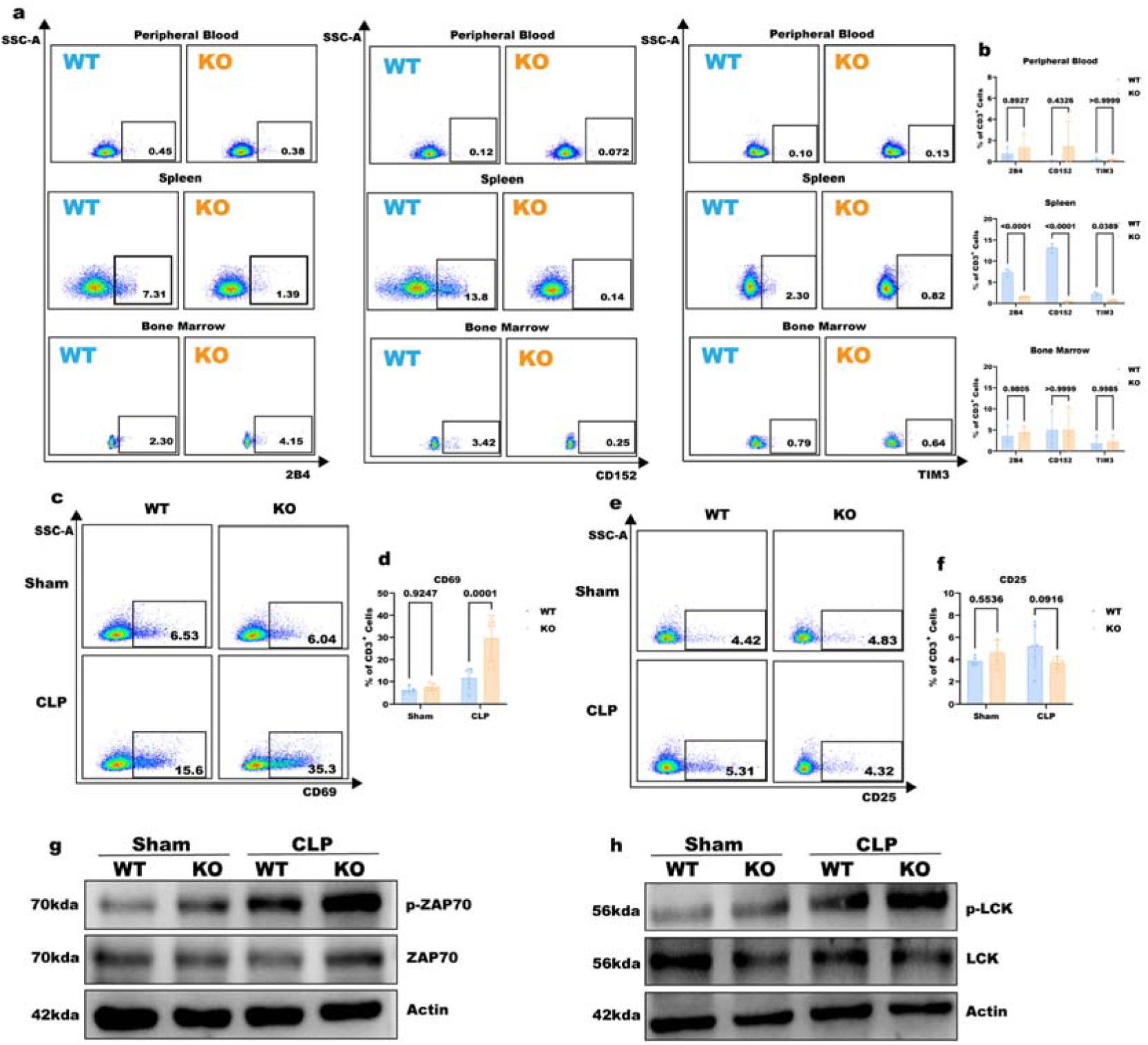
*Samsn1*^-/-^ mice have decreased T cell exhaustion-related receptors expression and enhanced T cell activation and signaling. **(a)** Flow cytometry showed the expression of 2B4, CD152, and TIM3 in T cells from peripheral blood, spleen, and bone marrow of WT and *Samsn1*^-/-^ mice. **(b)** The changes in the proportion of CD3^+^ T cells in WT and *Samsn1^-/-^*mice were quantified. **(c, e)** Flow cytometry analysis of CD69 and CD25 expression in spleen T cells of WT and *Samsn1^-/-^* mice 12 hours after CLP. **(d, f)** The changes in the proportion of CD3^+^ T cells were quantified. **(g, h)** WB analysis showed the phosphorylation of ZAP70 and LCK in spleen T cells from WT and *Samsn1^-/-^* mice after Sham or CLP surgery. Statistical analysis of **(b, d, f)** was performed using *2-way ANOVA test* (n=5).

### SAMSN1 binds to KEAP1, releasing NRF2 from KEAP1 and allowing NRF2 to enter the nucleus to promote the transcription of target genes CD48, CD86 and CEACAM1

Finally, we explored how SAMSN1 affects the expression of CD48, CD86, and CECAM1 in macrophages. We first analyzed the composition of the protein complexes bound to SAMSN1 using IP-MS (immunoprecipitation of proteins bound to SAMSN1 followed by mass spectrometry). We identified 81 and 74 proteins interacting with SAMSN1 in resting RAW264.7 cells (WT-REST) and LPS-stimulated RAW264.7 cells (WT-LPS), of which 38 genes were shared by both groups, 43 were unique to WT-REST, and 36 were unique to WT-LPS (raw data of mass spectrometry analysis are shown in Supplementary Materials 2). We speculated that if SAMSN1 regulates the expression of CD48, CD86 and CEACAM1 through some pathway, it may be achieved by binding to a certain transcription factor. We analyzed the 43 genes unique to WT-REST and the 36 genes unique to WT-LPS to see if any transcription factors were present. Among them, there are 7 transcription factors that bind to SAMSN1, and 4 of them (JUNB, ZFP36, NFKIBZ, and KEAP1) can be found in public databases for transcriptome sequencing data after knocking them out in macrophages (Fig. 8a). The 4 public datasets GSE50542, GSE229923, GSE43075, and GSE216252 show the RNA-Seq data of macrophages after JUNB, ZFP36, NFKIBZ, and KEAP1 were knocked out, respectively. We analyzed these datasets and found that the expression of CD48, CD86 and CEACAM1 was significantly reduced after KEAP1 knockout, while the deletion of JUNB, ZFP36, NFKIBZ genes had no effect on the expression of these genes (Fig. 8b and Supplementary Fig. 15). The results suggest that SAMSN1 may regulate the expression of CD48, CD86 and CEACAM1 by binding to the transcription factor KEAP1.

**Figure 8.**
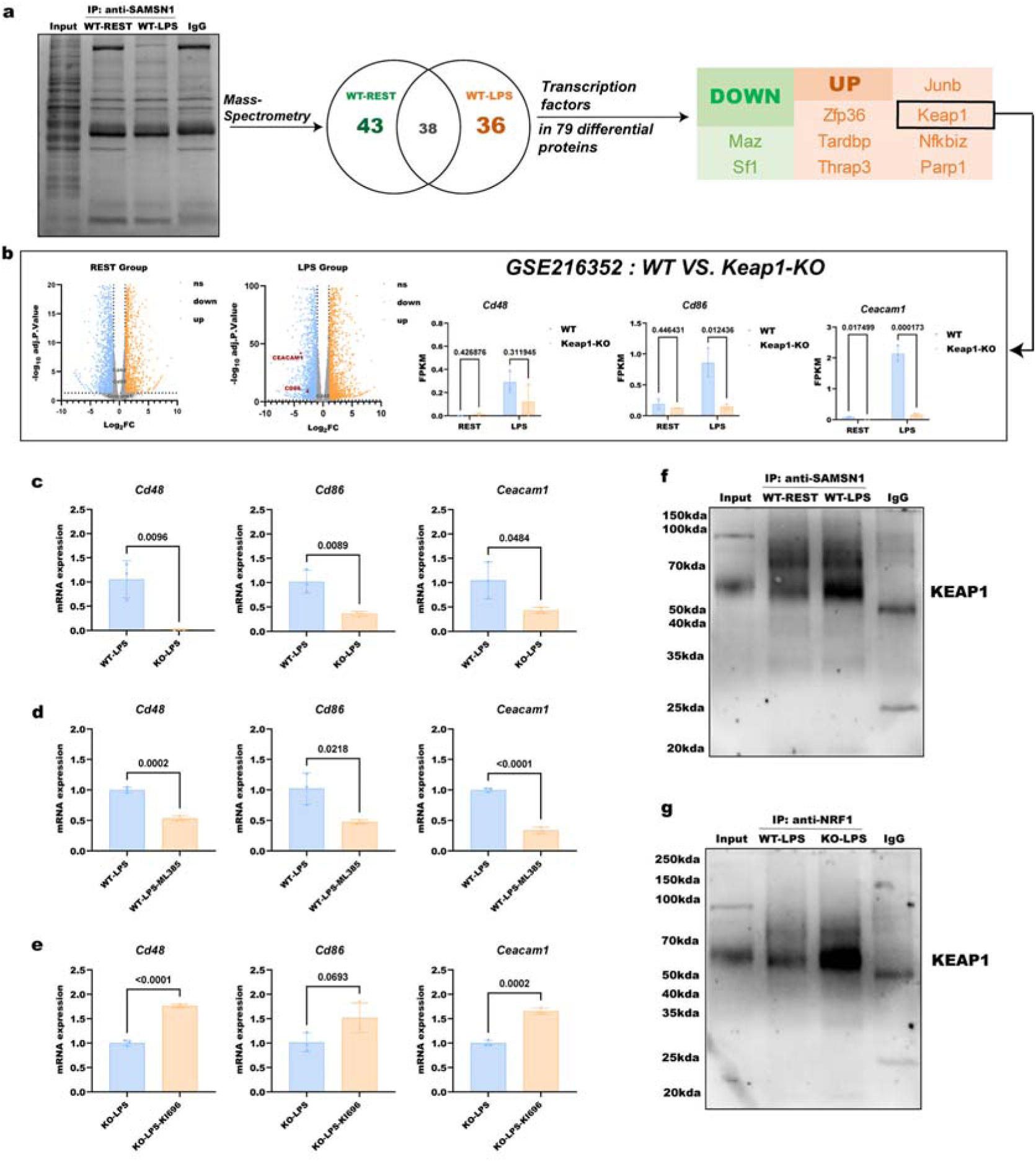
Identification of regulatory factors that bind to SAMSN1 and regulate transcription of downstream genes. **(a)** The flow chart shows that proteins bound to SAMSN1 are first precipitated by IP, and then the transcription factors that bind to SAMSN1 and change under LPS stimulation are analyzed by mass spectrometry. Among the transcription factors that are upregulated after LPS stimulation, JUNB, ZFP36, NFKIBZ and KEAP1 can be found in the public datasets for RNA-Seq data after knocking them out. **(b)** GSE216352 dataset shows the expression of co-inhibitory molecules CD48, CD86, and CEACAM1 in WT and KEAP1-KO RAW264.7 cells. **(c)** Real-time RT-PCR detection of CD48/CD86/CEACAM1 mRNA levels after WT or SAMSN1-KO cells were stimulated with LPS. **(d)** Real-time RT-PCR detection of CD48/CD86/CEACAM1 mRNA levels in WT cells stimulated with LPS after treatment with the KEAP1 inhibitor ML385 (5μM for 1h). **(e)** Real-time RT-PCR detection of CD48/CD86/CEACAM1 mRNA levels in SAMSN1-KO cells stimulated with LPS after treatment with the agonist KI696 (20μM KI696 for 24h). **(f)** IP-WB assay showed that the specific binding of SAMSN1 and KEAP1 increased when WT cells were stimulated with LPS. **(g)** IP-WB assay detected the binding of KEAP1 and NRF2 in LPS-stimulated WT or SAMSN1-KO cells. Significance in **(b-e)** was assessed by *2-tailed Welch’s t test*.

Nuclear factor erythroid 2-related factor 2 (NRF2) plays pleiotropic roles in a variety of physiological and pathological regulation. In the resting state, KEAP1 binds to NRF2 in the cytoplasm, preventing NRF2 from translocating into the nucleus to trigger the transcription of target genes^13–15^. IP-MS analysis has revealed that SAMSN1 and KEAP1 bind to each other upon LPS stimulation. We hypothesized that in the resting state, KEAP1 and NRF2 are bound in the cytoplasm, and SAMSN1 and KEAP1 are separated. However, under the stimulation of pathogenic components such as LPS, SAMSN1 binds to KEAP1 and promotes the release of NRF2 from KEAP1, so that NRF2 translocates into the nucleus and promotes the transcription of CD48, CD86 and CEACAM1.

We used two agents, ML385, which specifically inhibits NFR2, and KI696, which specifically blocks KEAP1 and NRF2 binding thereby activates NRF2, to test this hypothesis. Realtime-RT-PCR confirmed that the LPS-induced expression of CD48, CD86 and CEACAM1 could be significantly reduced when SAMSN1 gene was deleted (Fig. 8c). ML385 significantly inhibited the LPS-induced expression of CD48, CD86 and CEACAM1 (Fig. 8d). On the other hand, under the stimulation of LPS, the use of agonist KI696 can significantly restore the expression of CD48/CD86/CEACAM1in SAMSN1-KO cells (Fig. 8e). The results suggest that NRF2 is a key downstream regulator of SAMSN1 to promote the transcription of the co-inhibitory molecules. IP-WB analysis showed that the specific binding of SAMSN1 and KEAP1 increased when WT cells were stimulated with LPS (Fig. 8f). However, in SAMSN1-KO cells, the binding of KEAP1 and NRF2 increased significantly under the stimulation of LPS, suggesting that the loss of SAMSN1 increased the number of NRF2 binding to KEAP1, thus inhibiting its transcriptional activity (Fig. 8g).

## Discussion

In order to screen out key genes that may be involved in the pathogenesis of sepsis, we retrieved a total of 56 public sequencing datasets and selected three datasets that met the screening criteria (GSE131761, GSE154918, and GSE139913), in which a gene named SAMSN1 was screened out. Screening of public datasets and external validation of prospective data revealed that SAMSN1 expression was significantly increased in patients with sepsis and was correlated with sepsis severity and mortality. In vivo experiments showed that SAMSN1 knockout or blockade with antibodies improved the survival of septic mice via enhancing macrophage proliferation, phagocytosis, and clearance of bacteria from the blood. SAMSN1 induces macrophages to be immunosuppressive by upregulating the immune co-inhibitory molecules CD48, CD86, and CEACAM1, which bind to the corresponding receptors 2B4, CTLA4, and TIM3 on the surface of T cells, leading to T cell exhaustion. Through IP-MS analysis, we identified that SAMSN1 binds to a transcription factor partner KEAP1, thereby dissociating the KEAP1-NRF2 complex in the cytoplasm, so that NRF2 translocates into the nucleus and initiates the transcription of downstream target genes CD48, CD86 and CEACAM1. The schematic of the molecular mechanism of SAMSN1 is shown in Fig. 9.

**Figure 9.**
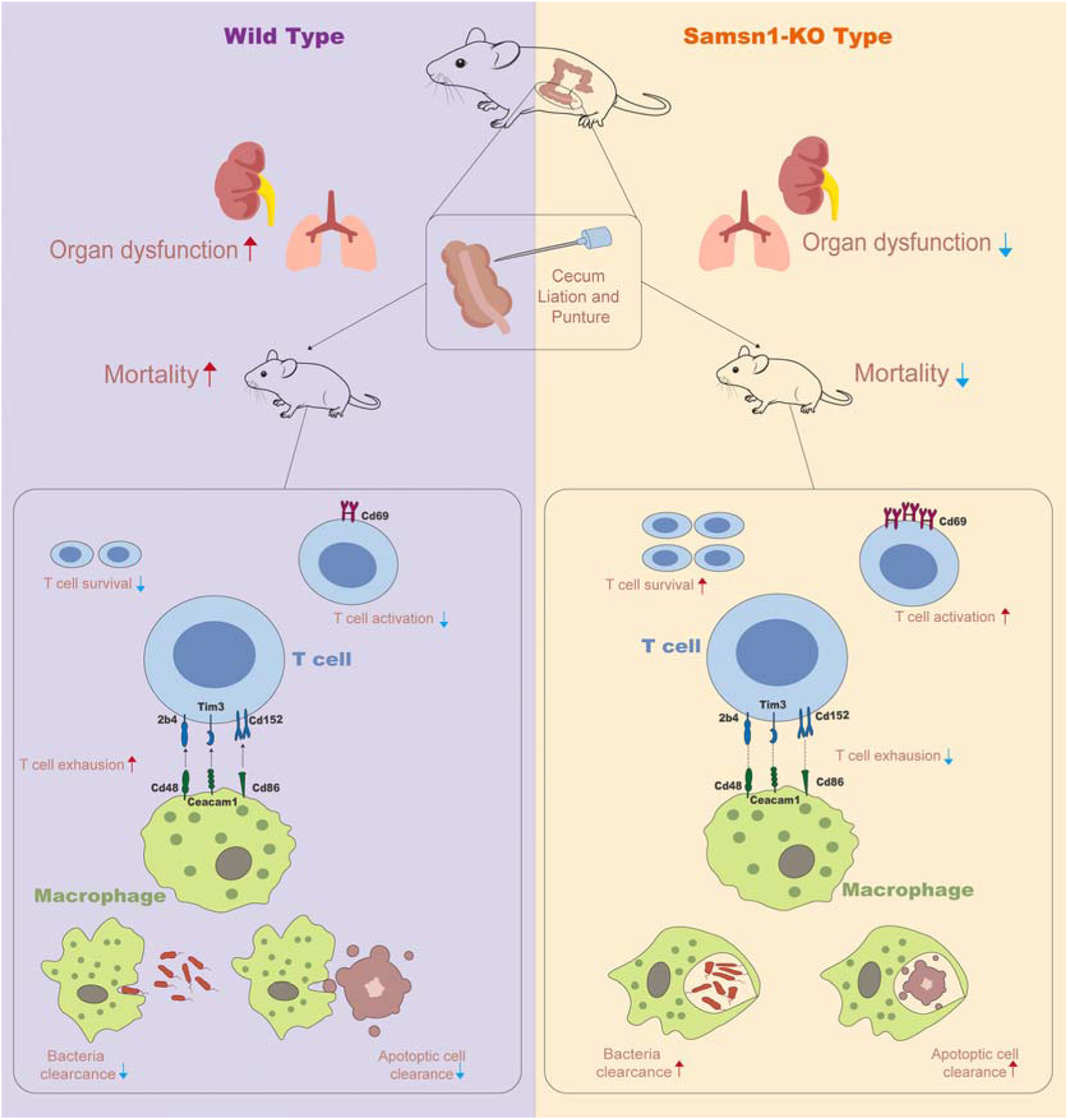
SAMSN1 affects macrophage-T cell cross-talk in sepsis immunosuppression. Schematic diagram: In the resting state, KEAP1 and NRF2 form a transcription complex in the cytoplasm of monocyte-macrophages and are in an inhibitory state. When sepsis occurs, the number of monocyte-macrophages increases significantly, in which SAMSN1 is highly expressed. SAMSN1 binds to KEAP1, causing NRF2 to dissociate from the complex and translocate into the nucleus, promoting the transcription of co-inhibitory molecules CD48/CD86/CEACAM1. Through cell-cell contact, these co-inhibitory molecules bind to corresponding receptors 2B4/TIM4/CD152 on the surface of T cells, causing T cell inhibition and exhaustion. High expression of SAMSN1 also switches macrophages into a suppressive phenotype, manifested by decreased phagocytic ability and ability to clear bacteria in the blood. Blockade of SAMSN1 by gene knockout or monoclonal antibody keeps KEAP1-NRF2 in a complex state, preventing NRF2 from entering the nucleus and promoting the transcription of downstream co-inhibitory molecules, thereby failing to inhibit T cell activation. Blockade of SAMSN1 also switches macrophages to a pro-immune phenotype with enhanced phagocytic ability, resulting in reduced organ injuries and increased survival rate.

SAMSN1 was first identified and reported simultaneously by Claudio ^16^ and Uchida ^17^ in 2001. Claudio et al. found that SAMSN1 was highly expressed in acute myeloid leukemia cells and multiple myeloma cells and named it HACS1 ^16^, whereas Uchida et al. identified SAMSN1 in mast cells and named it NASH1 ^17^. SAMSN1 has also been found to be highly expressed in other tumors ^18–23^. Another alias for SAMSN1 is SLy2, which, together with SLy1 and SLy3, belongs to the adaptor protein family SLy/SASH1 ^24^. The SLy family has a highly similar protein structure, including the SH3 motif, SAM1 domain, and nuclear localization signal (NLS). Earlier studies have suggested that SAMSN1 mainly functions on B cells ^25–29^. However, our analysis of the single-cell transcriptome sequencing dataset GSE151263 showed that SAMSN1 is expressed in many types of immune cells, with the highest expression in monocyte-macrophages. Wang et al. reported that conventional knockout of SAMSN1 resulted in overall immune enhancement in T cells, B cells, and DC cells of mice ^30^. Our data suggest that SAMSN1 can bind to the transcription factor partner KEAP1, and SAMSN1 knockout resulted in KEAP1-NRF2 to remain in a bound state, and NRF2 could not dissociate from KEAP1 and enter the nucleus, thus failing to activate the expression of immunosuppressive co-stimulatory molecules CD48, CD86, and CEACAM1. After SAMSN1 knockout, in addition to the to the increase in T cells, B220^+^CD5^+^ B cells also increased accordingly (Supplementary Fig. 9), suggesting that the recovery of T cell activity may also promote B cell function.

In the past few decades, all drugs designed to target inflammatory mediators have failed, suggesting that inflammation may not be the core of the pathogenesis of sepsis. The role of immunosuppression in the pathogenesis of sepsis has been recognized in recent years, but the mechanism that triggers immunosuppression in sepsis has not yet been fully elucidated. In this study, we proposed for the first time that SAMSN1 in monocytes-macrophages may mediate crosstalk between monocyte-macrophages and lymphocytes, causing T cell exhaustion, and SAMSN1 knockout can relieve immunosuppression and significantly improving survival in sepsis. According to the analysis of The Human Protein Atlas database, SAMSN1 is mainly located in the cell membrane and partly in the nuclear membrane. Therefore, it is theoretically feasible to use antibodies to inhibit SAMSN1 activity. We prepared 5 monoclonal antibody clones that can bind to SAMSN1. Among them, mAb-10-A2-H10 showed a significant improvement in survival rate in septic mice. Based on this, we may be able to design novel immune-boosting drugs for sepsis treatment.

Previous studies have shown that SAMSN1 is expressed and functions in B cells, mast cells or tumor cells. According to our analysis of single-cell sequencing data of PBMC, SAMSN1 is mainly expressed on monocyte-macrophages, which will broaden our understanding of the biological function of SAMSN1. There is currently a lack of reports on the relationship between SAMSN1 and sepsis and septic immunosuppression. To the best of our knowledge, we show for the first time that SAMSN1 expression is significantly increased during sepsis and exerts an immunosuppressive function. SAMSN1 induces an immunosuppressive phenotype in monocyte-macrophages by up-regulating co-inhibitory molecules CD48/CD86/CEACAM1, which bind to the corresponding receptors 2B4/CTLA4/TIM3 on the surface of T cells, leading to T cell exhaustion. Through proteomic identification by IP-MS, followed by transcriptome sequencing analysis, we identified the key molecules downstream of SAMSN1 regulation. SAMSN1 binds to KEAP1 to dissociate the KEAP1-NRF2 complex, allowing NRF2 to enter the nucleus and promote the transcription of its target gene immune co-suppressor molecules.

In this study, we first used public database to screen out genes that are significantly changed when sepsis occurs, and then performed external validation using a prospective cohort with RNA-Seq data, and finally used gene editing to study the function of the target gene and conduct sepsis therapeutic experiments. This stepwise screening process using a multi-omics approach improves the objectivity of the research and makes the findings more potentially translational. Our study proposes a novel mechanism for the triggering of immunosuppression in sepsis and may provide a new target for the diagnosis and treatment of sepsis.

## Methods

### Ethics

The observational study using human blood samples for RNA-Seq was approved by Ethics Committee on Biomedical Research, West China Hospital of Sichuan University (registered observational trial: ChiCTR2100047060). The informed consent was obtained from all subjects. This trial was examined and cleared by the Medical Ethics Committee under the ethical approval No.2020641.The informed consent was obtained from all subjects. The animal experiments were approved by the Animal Ethics Committee of Sichuan University and performed according to institutional and national guidelines.

### Cecal ligation and puncture (CLP)

WT C57BL/6 mice were purchased from GemPharmatech. *Samsn1^-/-^*mice were generated by GemPharmatech. All mice were caged at 22±L3L°C under 12h-12h light-dark cycles before cold and warm exposure, and air change rate of 12-15 times per hour are maintained at a constant level. The mice were randomly divided into groups before the experiment. Before CLP surgery, male mice (8-10 weeks, 20-25 g) were anesthetized with an intraperitoneal (i.p.) injection of Pentobarbital Sodium (50 mg/kg). An abdominal midline incision was performed and the caecum was isolated and ligated, then the caecum was ligated 7.5 mm from the caecal tip at a site distal to the ileocecal valve, and the ligated caecal stump was then perforated using two “through and through” punctures (18-gauge needle). The cecum was then returned to the peritoneal cavity, and the abdominal skin was closed. In the sham group, except for cecal ligation and puncture, other operation steps remained unchanged. All mice undergoing CLP surgery received antibiotics and fluid therapy, including a single intramuscular injection of Ciprofloxacin at a dose of 20 mg/kg and subcutaneous fluid resuscitation with 800 μl of saline immediately.

### Quantitative real-time PCR

Total RNA was isolated from samples using Trizol Reagent, which was obtained from Invitrogen. Reverse transcription was performed with a Superscript II Two-Step RT-PCR Kit (Invitrogen). PCR was performed using SYBR® Green PCR Master Mix (Applied Biosystems). Gene expression was normalized to 18s. The reaction mixture was preheated for 10 min at 95 °C, followed by 45 cycles of denaturation at 95 °C for 30 s, annealing at 55 °C for 30 s, and extension at 72 °C for 30 s. The 2−ΔΔCt method was employed to evaluate mRNA expression.

### ELISA

Serum was collected from mice at 8h, 1d, 3d, 5d, and 7d after CLP surgery. Samples collected for the measurement of IL-6, IL-10 and TNF-a levels with ELISA kits (R&D Systems) according to the manufacturer’s instructions.

### Cell lines and Crispr/cas9-mediated Samsn1 editing

RAW264.7 cells, HEK293 cells and jurkat cells were obtained from ATCC; RAW264.7 cells and HEK293 cells routinely cultured in complete DMEM, while jurkat cells cultured in complete RPMI-1640. All cells cultured at 37 °C in a 5% CO_2_ incubator. We designed sgRNAs targeting Samsn1 exon and integrated it into lentiCRISPRv2-mcherry plasmid backbone (Addgene); then lentivirus was packaged by HEK 293 cells, after that RAW264.7 cells were infected using lentivirus; after screening by puromycin, flow cytometry sorting was used to spread cells into 96-well plate to obtain single cells; after culturing the monoclonal cells, the identification was performed to obtain the final *Samsn1^-/-^* RAW264.7 cell line.

### Survival study

For survival study-1: Male C57BL/6 WT mice were randomly assigned to the Sham (WT-Sham) and CLP (WT-CLP) group; Male C57BL/6 *Samsn1^-/-^* mice were KO-CLP group. For survival study-2: Male C57BL/6 WT mice were randomly assigned to the Sham, CLP, CLP+PBS, CLP+mAb-7-A7-H3, CLP+mAb-10-A2-H10, CLP+mAb-38-C5-C10, CLP+mAb-41-B2-H5 and CLP+ mAb-53-G11-H1 group. All 5 mAbs were purchased from AtaGenix. Mice were subjected to Sham or CLP surgery and excluded if they died during surgery. The survival of mice was monitored for up to 14 days.

### H&E staining and histologic scoring

The septic mice were anaesthetized and perfused transcardially with 4% paraformaldehyde in PBS for 10 min. The liver, lung and kidney were removed, post-fixed with 4% paraformaldehyde for 24 h, embedded in paraffin, and sectioned at a 5-μm thickness. After deparaffinization and rehydration, thesections were stained with hematoxylin and eosin. Based on the scoring standard made by Ferrer et al^31^. The H&E staining was scored by two blinded observers (See Supplementary Table 5).

### Cell proliferation

The cell counting plate was used to count the number of cells, and then the 96-well plate was inoculated with the same number of cells (5000 cells/well) and incubated for 2 h to wait for the cells to attach to the wall, then 50 ug/ml LPS was added to the medium; at 0, 1, 6, 12, 24, 48 and 72 hours, 10ul CCK-8 (MedChemExpress) was added to the medium and incubated for 1 h to check the OD value. The formula of Cell proliferation rate follows product protocol.

### Blood bacterial load

Blood samples were collected from the septic mice (WT and *Samsn1^−/−^*) 12 h after the CLP surgery. Ten microliters from each sample were plated on blood agar plates, and bacterial colonies (CFU) were counted manually.

### Bacterial phagocytosis assay

Escherichia coli (K-12 strain) BioParticles® conjugated to Alexa Fluor®488 (Invitrogen) were used according to the manufacturer’s instructions. RAW264.7-WT and RAW264.7-KO Cells were seeded on 24-well plates and incubated for 1 h at 37 °C; the medium was then removed and replaced with medium treated with or without E. coli particles (1 mg/ml), and cells were incubated at 37 °C for 2 h. The phagocytic activity was determined by measuring the fluorescent signal within the macrophages using Image-Pro Plus software.

### Efferocytosis assay and Isolation of PPMs

Efferocytosis, phagocytosis of apoptotic cells. Construction of jurkat cells with green fluorescence: Calcein AM solution 1mg/ml (beyotime) was added to jurkat cells and incubated for 2h. Construction of ACs: Sraurosporine 1uM was added to jurkat cells (MedChemExpress) was incubated and then the degree of apoptosis was detected by flow cytometric staining for Annexin V/PI. Isolation of PPMs: Mouse peritoneal cells were obtained by repeated lavage of the mouse peritoneal cavity, incubated overnight to make them adherent to the wall, and the supernatant was removed to obtain PPMs. The prepared ACs and PPMs were co cultured in 1:1 ratio for 0,30 and 90 minutes.

### Flow cytometry and antibodies

Antibodies to CD3 (Clone: 17A2, Catalog: 100204), CD4 (Clone: GK1.5, Catalog: 100449/100408), CD8a (Clone: S18018E, Catalog: 162306), B220 (Clone: RA3-6B2, Catalog: 103234), CD5 (Clone: 53-7.3, Catalog: 100606), CD11b (Clone: M1/70, Catalog: 101212), CD152/CTLA-4 (Clone: UC10-4B9, Catalog: 106306), CD244.2/2B4 (Clone: m2B4 (B6)458.1, Catalog: 133512), CD366/TIM3 (Clone: RMT3-23, Catalog: 119721), CD48 (Clone: HM48-1, Catalog: 103443), CD86 (Clone: PO3, Catalog: 105123), CD66a/CEACAM1 (Clone: MAb-CC1, Catalog: 134510), CD69 (Clone: H1.2F3, Catalog: 104512) and CD25 (Clone: A18246A, Catalog: 113704), were purchased from Biolegend. The fixable viability dye Zombie NIR™ Fixable Viability Kit and Zombie Violet™ Fixable Viability Kit were also purchased from Biolegend. Peripheral blood, spleen and bone marrow of WT and KO mice after CLP surgery were sampled for flow cytometry. Sampling was done 12h after CLP surgery. These tissues were homogenized by repeated pipetting and filtered through a 70 μm cell strainer, and then washed once with complete RPMI to prepare a single-cell suspension. The cells were stained with different markers according to different experimental protocols for 20 min in the dark. The expression of cell surface markers was analyzed using a BD LSRFortessa flow cytometer (BD Biosciences, USA). The data were analyzed using FlowJo software (FlowJo, Ashland, OR, USA).

### Isolation of Mouse splenic CD3^+^ cells and Co-culture

Mice were euthanized using carbon dioxide and immersed in 75% alcohol for 5 min; spleens were aseptically removed; then Grind the spleen and filtered through a 70um filter to obtain a single-cell suspension; Red blood cells were lysed using Red Blood Cell Lysis Buffer (Abcam); the concentration of cells was adjusted to 10^8^/ml; 100ul of cell suspension was taken and 100ul of Biotin-Antibody Cocktail (Biolegend) mixed and incubated on ice for 15 min; after washing with MojoSort^TM^ buffer (Biolegend), 10ul CD3^-^ Streptavidin Nanobeads (Biolegend) were added and incubated on ice for 15 min; 5 ml of buffer was added and the tubes were placed in MojoSort^TM^ magnet (Biolegend); unlabeled cells in the tubes were obtained after 5 min, and the washing was repeated 2-3 times, each time obtaining unlabeled cells in the tubes. Co-cultured RAW264.7 cells with wild-type primary T cells by direct contact way or indirect contact way. The co-culture ratio of RAW264.7 cells to primary T cells was 1:1. Precise counting ensured that the number of primary T cells in each well was equal; after 12h of co-culture, the remaining primary T cells were stained and counted precisely. Direct contact: Co-culture RAW264.7 and primary T cells directly; Indirect contact: using a 0.04um pore-size transwell to keep the two separated.

### RNA-Seq

Total RNA was isolated from WT-RAW264.7 cells and KO-RAW264.7 cells using Trizol Reagent. RNAs were delivered to Novogene Inc (Beijing, China). After total RNA qualification, mRNA enrichment, double-stranded cDNA synthesis, end repair poly-A&adptor addition, fragments selection and PCR, library quality assessment, illumina sequencing and quality control, we acquired RAW sequencing data and performed data analysis on it.

### Western blot

Cultured cells in 6-well plates and 100mg spleen from mice were harvested, washed twice with PBS, and lysed for 30 min on ice in 150 μl of RIPA Lysis Buffer (Beyotime) containing 1 mM PMSF (Beyotime) and Protease inhibitor cocktail (Beyotime). Proteins were quantified with the BCA method. Protein samples were resolved by 10% SDS-PAGE, transferred to a 0.45um PVDF membrane (Millipore), and blocked for 1 h with 5% BSA (Sigma-Aldrich) in Tris-buffered saline containing 0.1%Tween 20 (TBST) at room temperature. After incubation with a specific primary antibody overnight at 4 °C, the membrane was washed with TBST three times. Then, the membrane was incubated with a secondary antibody for 1 h at room temperature. Enhanced chemiluminescence (ECL) (Millipore) was used to visualize the protein bands, and images were acquired in a ChemiDoc™ Imaging System (Bio-Rad, Hercules, CA, USA). Each experiment was repeated at least three times. Primary antibodies: Anti-p-ZAP70 antibody (CST Cat. # 2717); anti-ZAP70 antibody (CST Cat. # 2705); anti-p-LCK antibody (CST Cat. #2751); anti-LCK antibody (CST Cat. # 2984); anti-Actin antibody (HUABIO); anti-Tubulin antibody (HUABIO); KEAP1; anti-SAMSN1 antibody (AtaGenix).

### Immunoprecipitation-Mass Spectrometry (IP-MS) and CO-IP

Anti-SAMSN1 Antibody (AtaGenix) and Anti-NRF2 Antibody (CST Cat. # 12721S) were used for the immunoprecipitation process of proteins from RAW264.7 cells. After RAW264.7 cells were grown to full size in 10mm cell dishes, with or without 50ug/ml LPS (Sigma) for 1h, then added to the NP-40 lysate for cell lysis, after which the supernatant was extracted by centrifugation at 4 °C 12,000g for 15 min, the collected supernatant was determined by the BCA method and taken for immunoprecipitation. Protein A/G magnetic beads (Selleck) were used to pre-mix with the antibody and incubated for 4h in a tumbler at 4 °C to obtain the magnetic bead-antibody complex. The magnetic bead-antibody complexes were co-incubated with prepared proteins and incubated overnight at 4 °C in a tumbler. According to the experimental design, the magnetic bead-antibody-protein complexes were sent to QLBio Inc (Beijing, China) for mass spectrometry analysis or western blot after denaturation to detect the expression of proteins.

### Statistical analysis

Data are presented as the means ± standard deviations (SDs) from at least three individual experiments. When two groups were compared, each set of data was tested for Normal (Gaussian) distribution first, and *Welch’s t test* (passed normality test) or *Mann-Whitney U test* (did not pass normality test) was used. For multi-group comparison, *one-way ANOVA test* followed by *Tukey’s multiple comparison* was used, and normality of Residuals was tested using “Anderson-Darling (A2*)”, “D’Agostino-Pearson omnibus (K2)”, “Shapiro-Wilk (W)”, and “Kolmogorov-Smirnov (distance)”. A p value < 0.05 was considered statistically significant. Survival outcomes were analysed using Kaplan-Meier survival curves, and the significance of the differences was assessed using the log-rank test.

## Data Availability

The RNA-Seq data used to validate the SAMSN1 mRNA levels in Fig. 1c, d have been deposited in the Genome Sequence Archive (Genomics, Proteomics & Bioinformatics 2021) in National Genomics Data Center (Nucleic Acids Res 2022), China National Center for Bioinformation/ Beijing Institute of Genomics, Chinese Academy of Sciences (GSA-Human: HRA002988) that are publicly accessible at https://ngdc.cncb.ac.cn/gsa-human. The RNA-Seq data of RAW264.7-WT and RAW264.7-KO cells in Fig. 6 have been deposited in the NCBI/SRA repository, accession number GSE262688 (https://www.ncbi.nlm.nih.gov/geo/query/acc.cgi?acc=GSE262688). All experimental data and detailed information about this study should be directed to Prof. Wei Zhang.

https://ngdc.cncb.ac.cn/gsa-human/browse/HRA002988

