## Supplementary file for "SAMSN1 causes sepsis immunosuppression by inducing macrophages to express coinhibitory molecules that causes T cell exhaustion via KEAP1-NRF2 signaling"

**Table S1**

| Search query | ("Homo sapiens"[Organism] AND ("sepsis"[Title] OR "septic"[Title]) AND ("2016/01/01"[PDAT] : "2022/01/01"[PDAT])) AND ("Expression profiling by array"[Filter] OR "Expression profiling by high throughput sequencing"[Filter]) | **56** |
| --- | --- | --- |
| Exclusion(53) | Not sepsis | **3** |
|  | Pediatric sepsis | **5** |
|  | Does not meet Sepsis 3.0 criteria | **9** |
|  | No health control | **26** |
|  | The sequencing subjects is not of interest(brain,heart,THP-1 and primary cells treated in vivo) | **6** |
|  | Secondary analysis or unable to analyzed | **4** |
| Inclusion(3) | GSE154918 | **3** |
|  | GSE131761 |  |
|  | GSE139913 |  |

**Search strategy in the GEO database.** Based on the search strategy, 49 of the 53 retrieved datasets were excluded from the search, resulting in the inclusion of 3 datasets to be analyzed

**Table S2**

| Series Accession | Sample | Groups | |
| --- | --- | --- | --- |
|  |  | Sepsis | Health Control |
| GSE154918 | Peripheral blood | 20 | 40 |
| GSE131761 | Peripheral blood | 81 septic shock | 15 |
| GSE139913 | circulating CD14+ monocytes | 4 | 5 |

**Details of the three selected dataset.** Sample types and groups for GSE154918,GSE131761 and GSE139913

**Table S3**

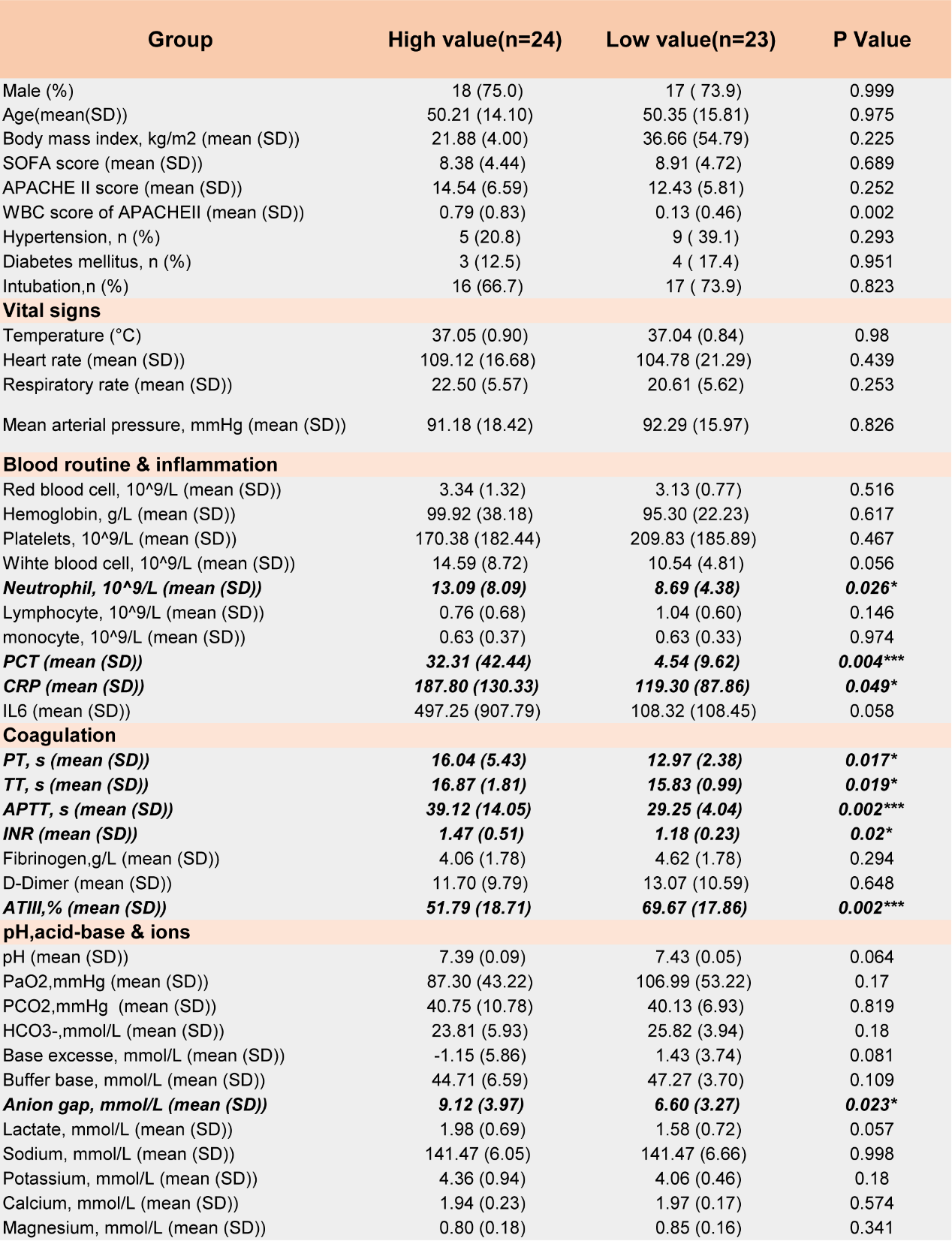

*(continued on next page)*

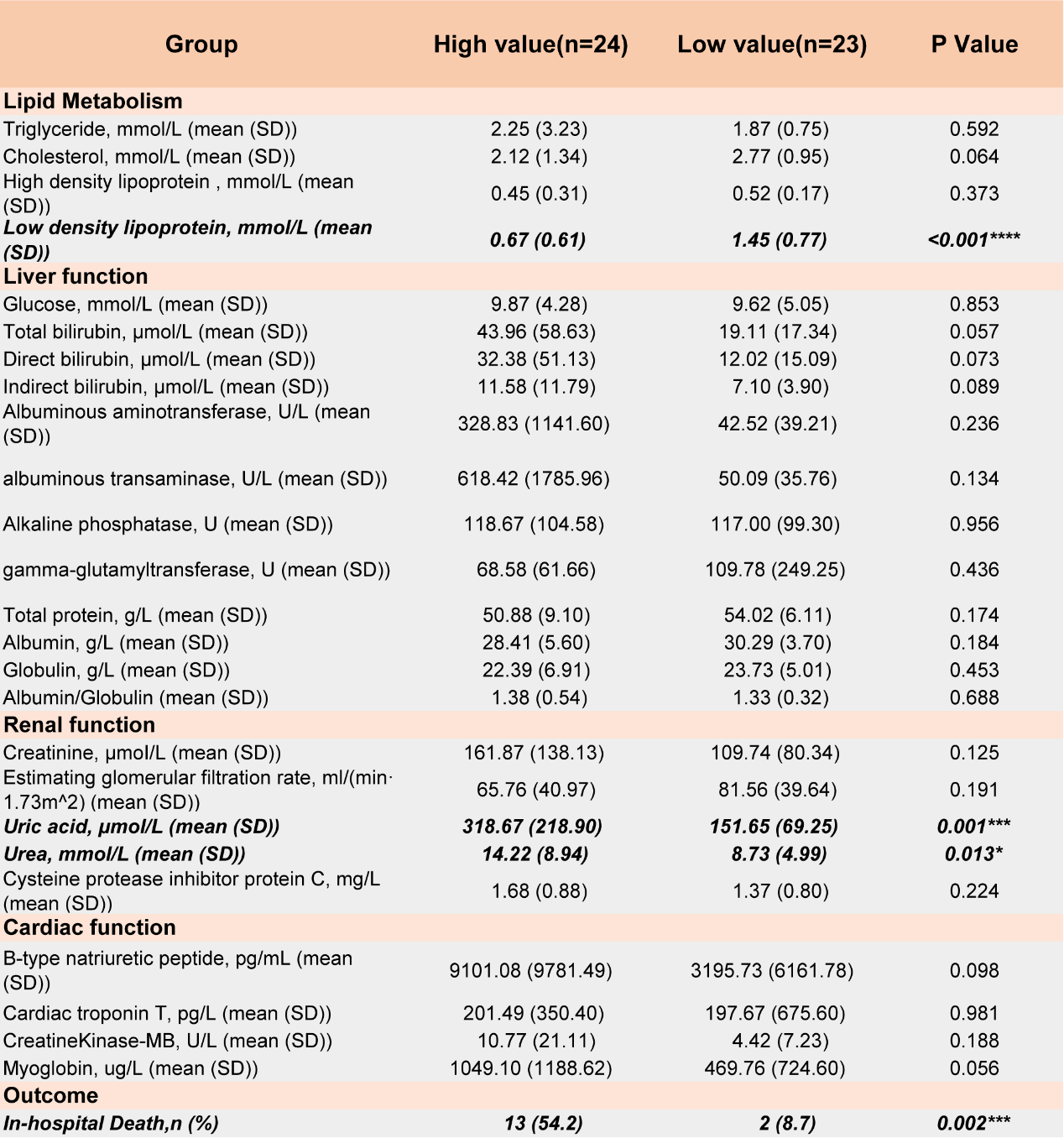

**Characteristics of septic patients with high or low value for *SAMSN1*.** PCT, procalcitonin; CRP, C-reactive protein; PT, Prothrombin time; TT, thrombin time ; APTT, Activated Partial Thromboplastin Time;, INR, International Normalized Ratio; ATIII, antithrombin Ⅲ.

**Table S4**

Histological Findings and Scoring Standard

| **Histological score** | **Organ** | | |
| --- | --- | --- | --- |
|  | **lung** | **liver** | **kidney** |
| 1 | Normal | Normal | Normal |
| 2 | Edema, congestion, **slightly** alveolar macrophages infiltration, **small area** hemorrhage | swollen hepatocyte, disappearance of sinus hepaticus, **slightly** periportal neutrophil and macrophage infiltration | Edema, congestion, **slight** leukocytes infiltration in glomeruli, vacuolar degeneration in the nucleus of epithelial cells |
| 3 | **Large area** hemorrhage, **severe** macrophage infiltration and proliferation, consolidation involving half of the lung | Changes seen above plus: lightly dyed nucleus or obscure nucleus of hepatocytes, vacuolar degeneration or ballooning degeneration of hepatocytes, **severe** periportal neutrophil and macrophage infiltration | Changes seen above plus: acute tubular atrophy, **severe** leukocytes infiltration in glomeruli, protein cast in the medulla, glomerulosclerosis |
| 4 | Changes seen above plus: **Severe** hemorrhage and consolidation involving virtually the whole lung | Changes seen above plus: granulomas with lipid laden macrophages, lymphoplasmocyter reaction, focal necrosis of hepatocyte |  |

**Fig. S1**

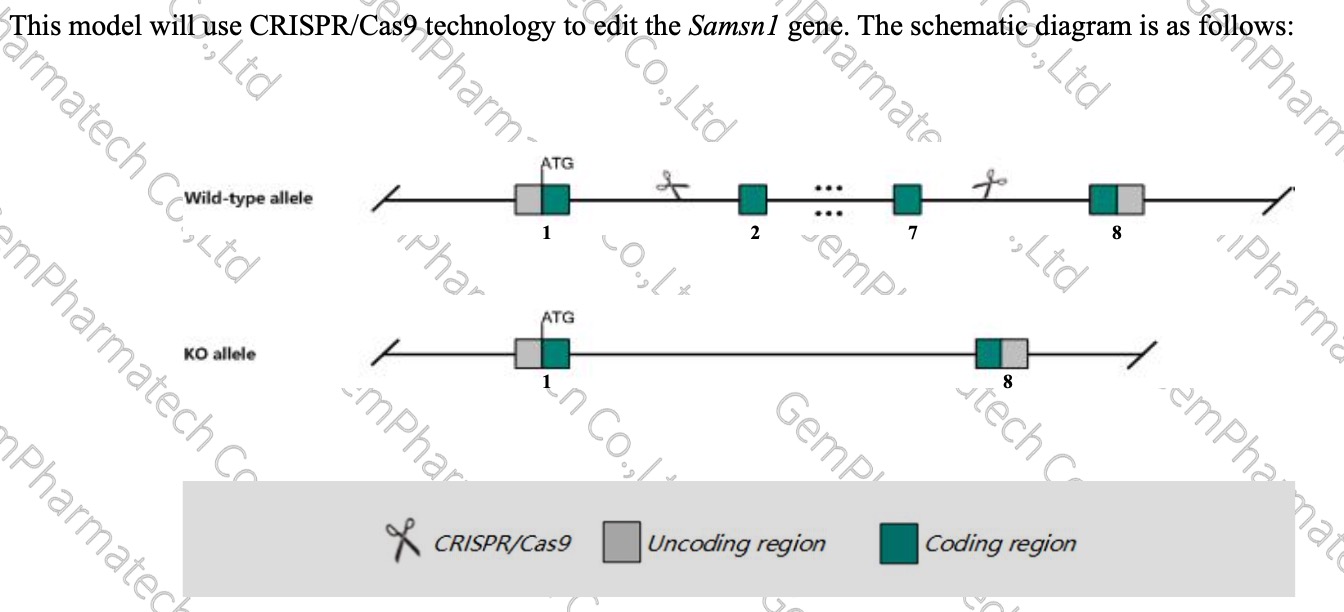

**Schematic diagram for editing *Samsn1* gene using CRISPR/Cas9.** The shear port is designed to shear the majority of the Samsn1 coding region.

**Fig. S2**

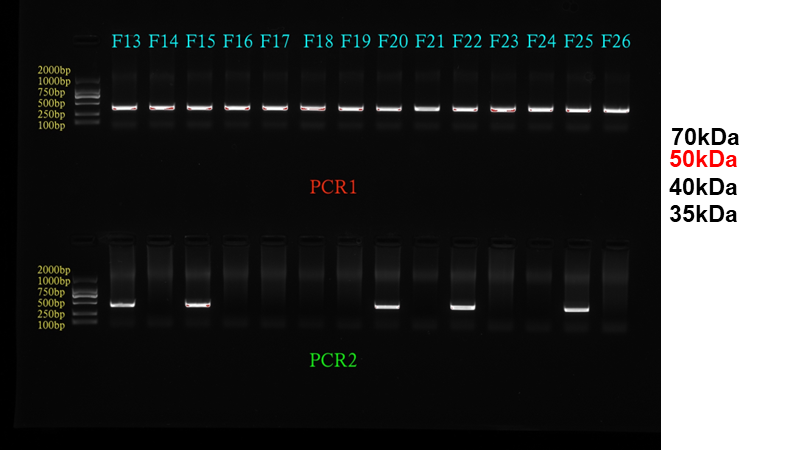

**Mouse tail genotype identification.** Samsn1 knockout mice created using Crispr/cas9 technology were bred to wild-type mice, followed by several generations of breeding to identify Samsn1 knockout purebred mice. The figure shows that PCR1 positive and PCR2 negative represent homozygotes; PCR1 positive and PCR2 positive represent heterozygotes; PCR1 negative and PCR2 positive represent wild-type.

**Fig. S3**

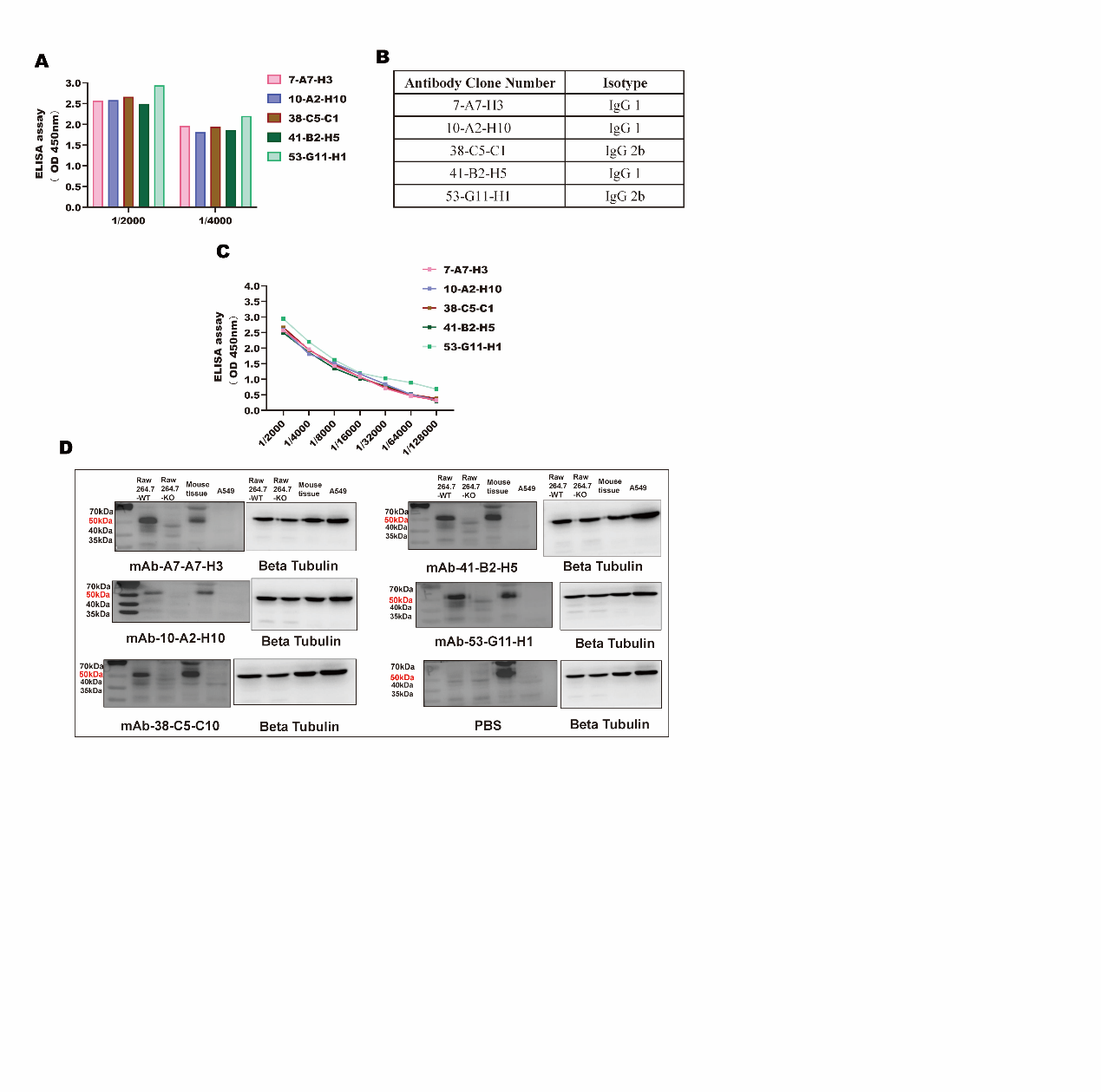

**The preparation of anti-SAMSN1 monoclonal antibodies.** (A) The binding of the antibody subclones to mouse-SAMSN1 at indicated concentrations was measured by ELISA assay(450nm). (B) The isotype identification showed that these five antibodies are mainly lgG2b and IgG 1. (C) The binding of these antibodies to mouse-SAMSN1 at the concentrations of 1/2000, 1/4000, 1/8000, 1/16000, 1/32000, 1/64000, 1/128000 were measured by ELISA assay(450nm). (D) Western blot for five antibodies. RAW264.7-WT, Wild type RAW264.7 cells protein; RAW264.7-KO, *Samsn1* knockout RAW264.7 cells protein; mouse tissue, mouse lung tissue protein; A549, A549 cells protein.

**Fig. S4**

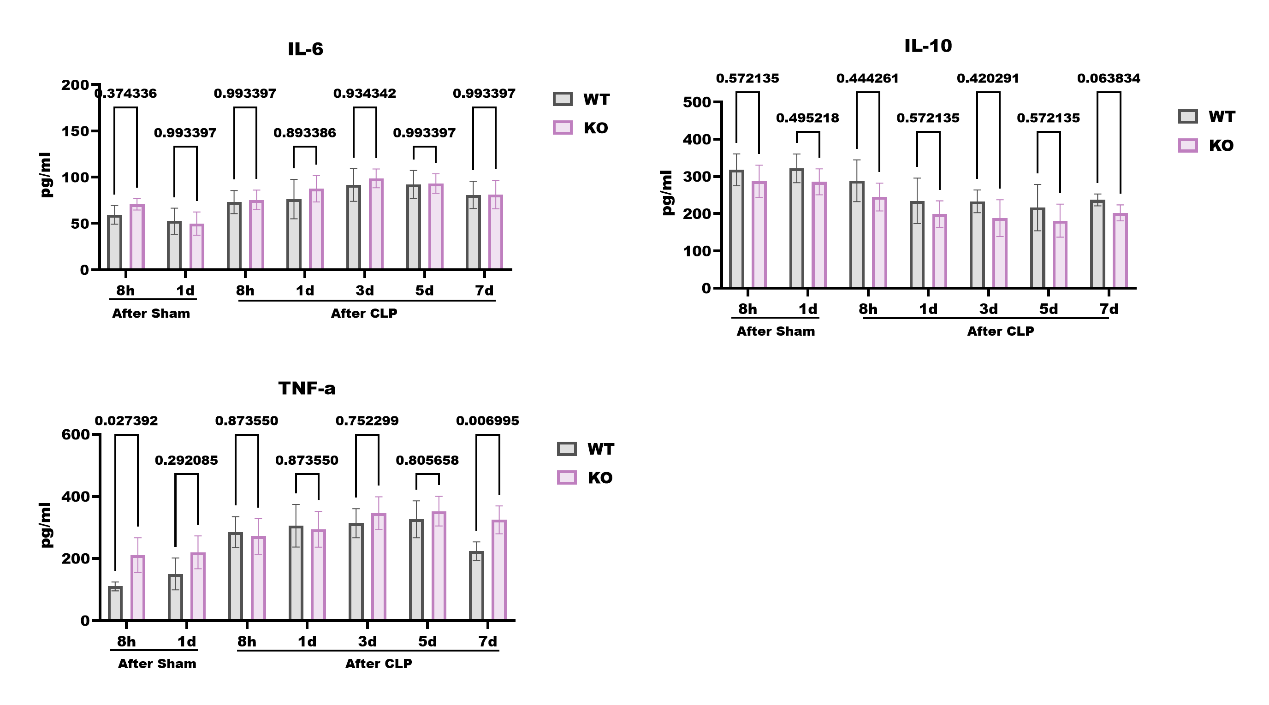

**Expression of IL6, IL0 and TNF-a in serum after CLP surgery.** Multiple unpaired t tests were performed to compare difference between WT and KO groups. (n=3-6 for each group). Data are shown as mean ± SD.

**Fig. S5**

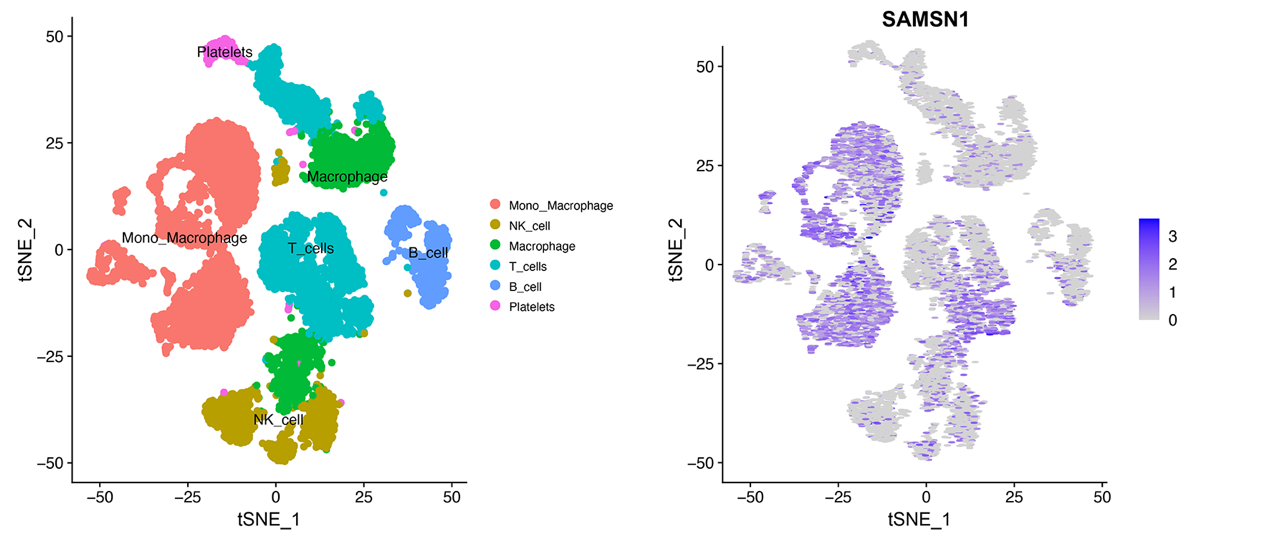

**The expression profile of SAMSN1 in human PBMC.** The expression of SAMSN1 in PBMCs of sepsis patients in the dataset GSE151263 and found that SAMSN1 was expressed on various types of immune cells, but mainly in monocytes-macrophages

**Fig. S6**

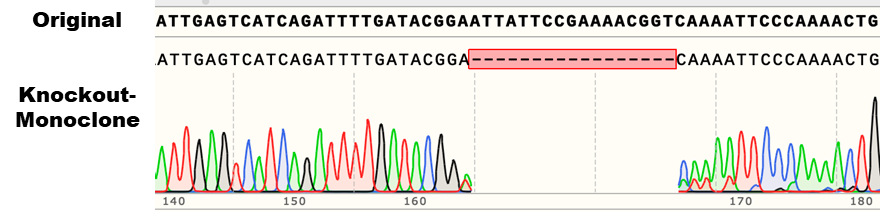

**Sanger sequencing of *Samsn1* knockout RAW264.7 cells.** Above, gene sequences of wild-type cell lines; below, gene sequences of *Samsn1* knockout monoclonal cell lines

**Fig. S7**

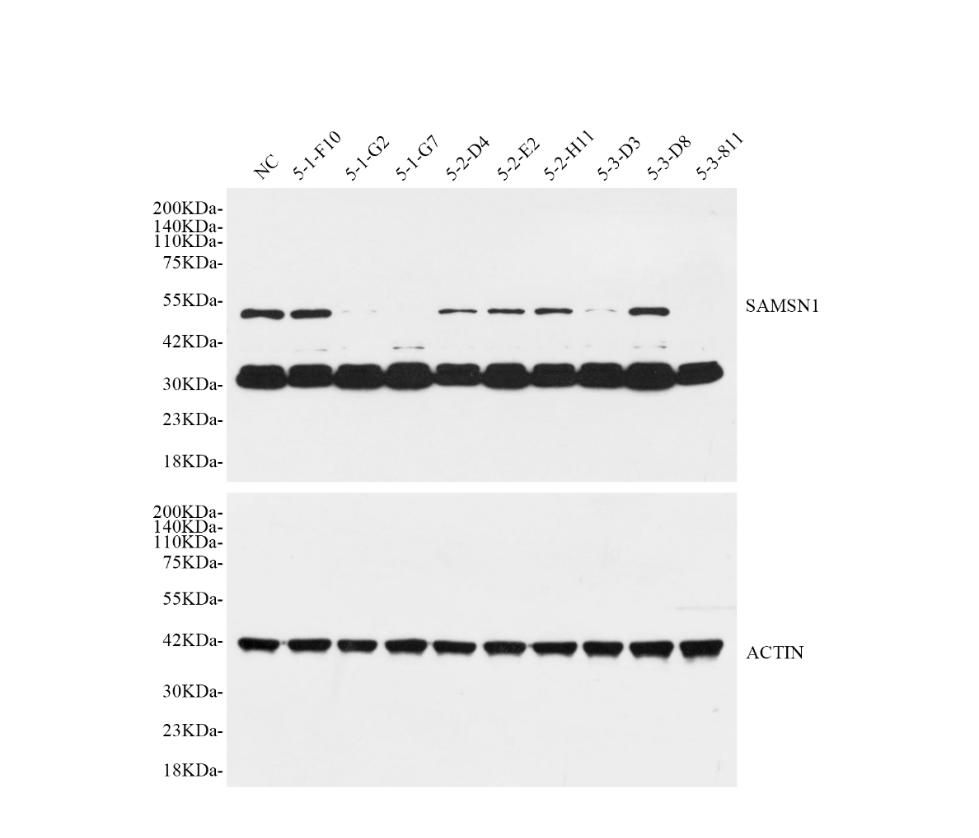

**Western blot identification of *Samsn1* knockout cell lines.**

**Fig. S8**

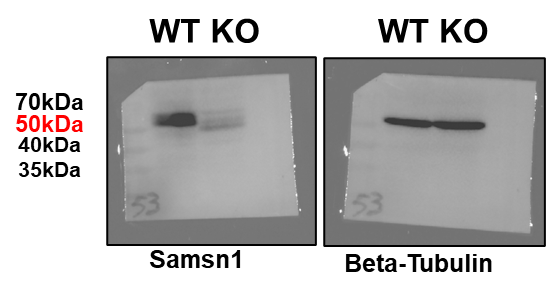

**Western blot identification of *Samsn1* knockout type C57BL/6 mice.**

**Fig. S9**

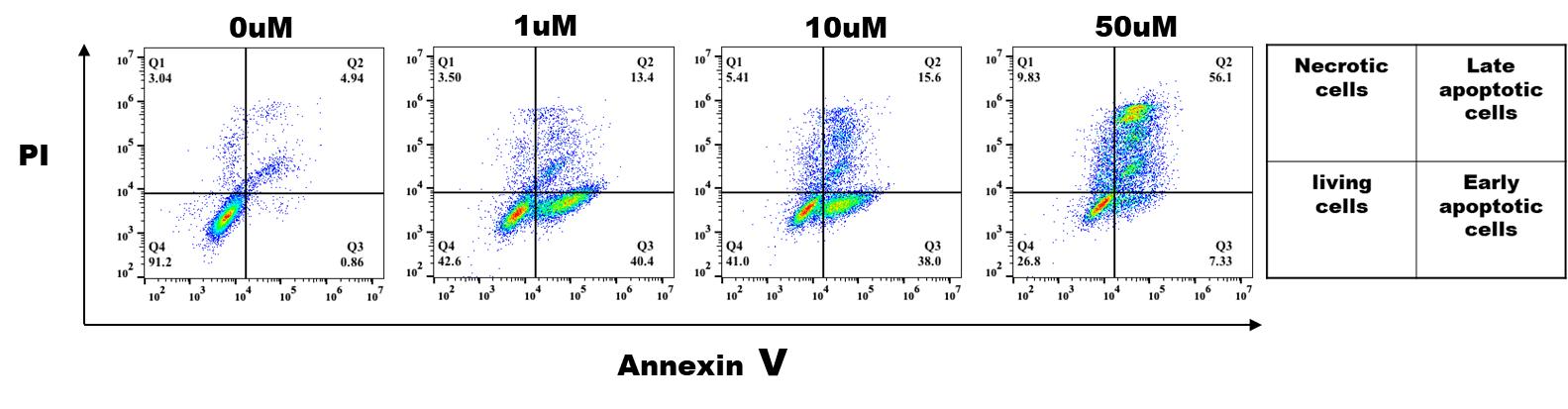

**Induction of apoptosis in Jurkat cells using different concentrations of Staurosporine.** Detection of the extent of apoptosis in jurkat cells using flow cytometry labeling with Annexin V/PI.

**Fig. S10**

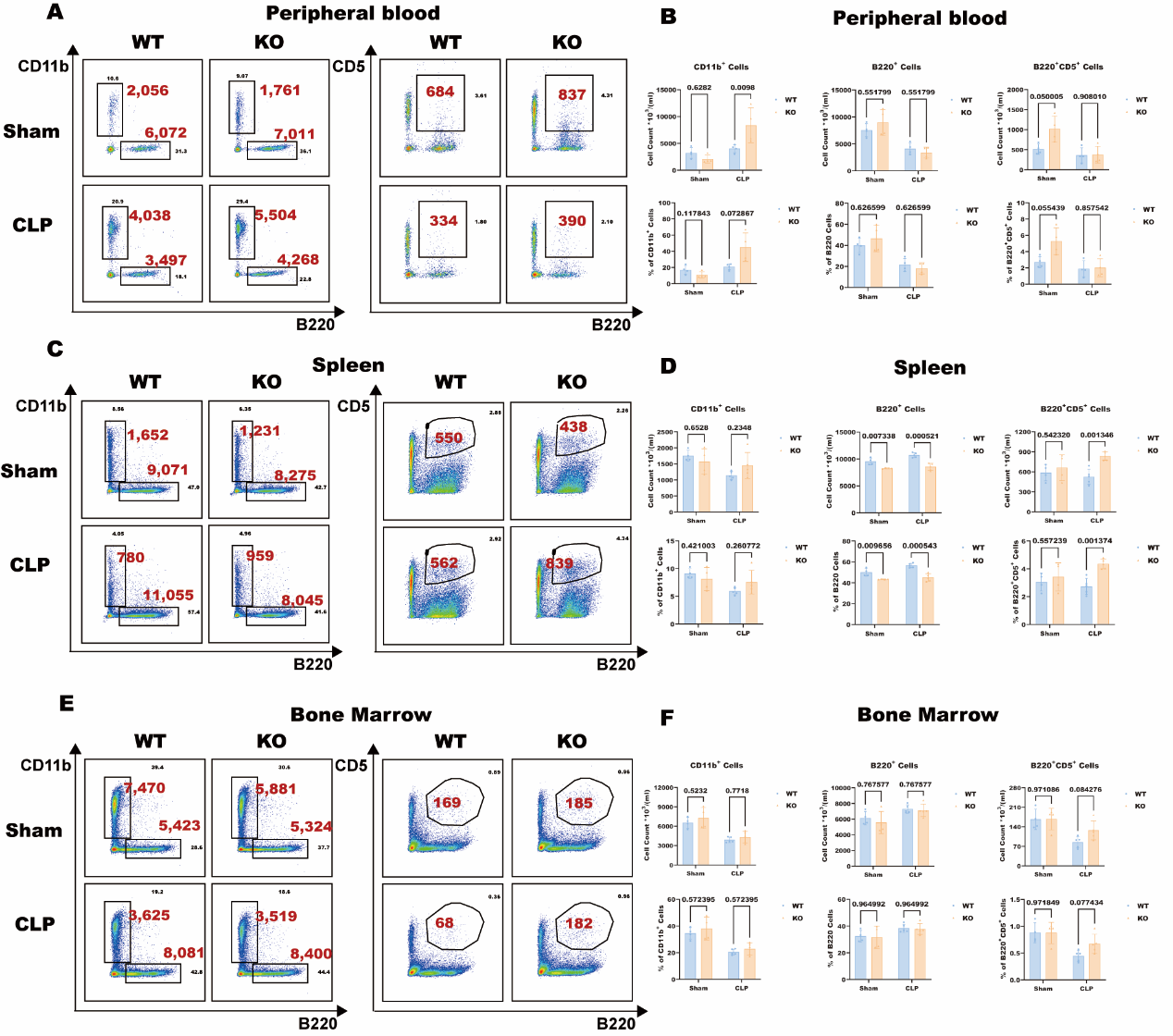

**B cells and CD11b cells after CLP surgery.** Detection of B cells (B220 and B220CD5 cells) and Cd11b cells in peripheral blood, spleen and bone marrow of WT and KO mice after CLP surgery by flow cytometry. Sampling was done 12h after CLP surgery. The graphs and histograms (A&B, peripheral blood; C&D, spleen; E&F, bone marrow) show the proportions of B220, B220CD5, and CD11b cells in the corresponding tissues of the mice as well as the number of cells calculated from the corresponding proportions (Red). Statistics was performed using multiple unpair t test. (n=3 for each group). Data are shown as mean ± SD in (B), (D) and (F).

**Fig. S11**

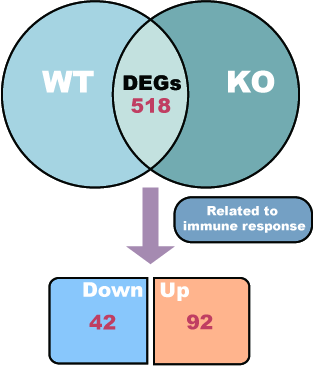

**Schematic for searching DEGs related to immune response.**

**Fig. S12**

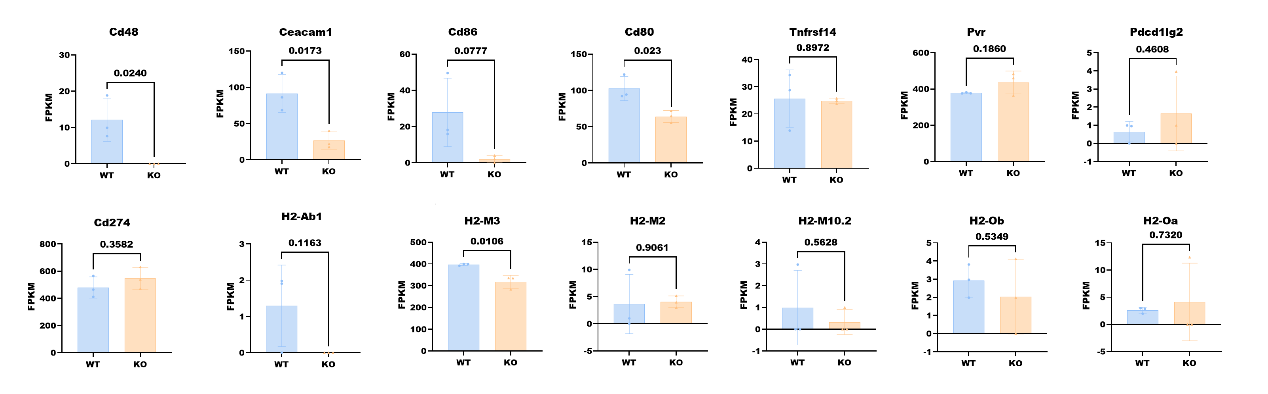

**Expression of ligands for T cell exhaustion-related receptors on RAW264.7 cells.** Data are shown as mean ± SD. Differences in expression (FPKM) between WT (n=3) and KO (n=3) were compared using the Unpaired t test or Mann-Whitney U-test.

**Fig. S13**

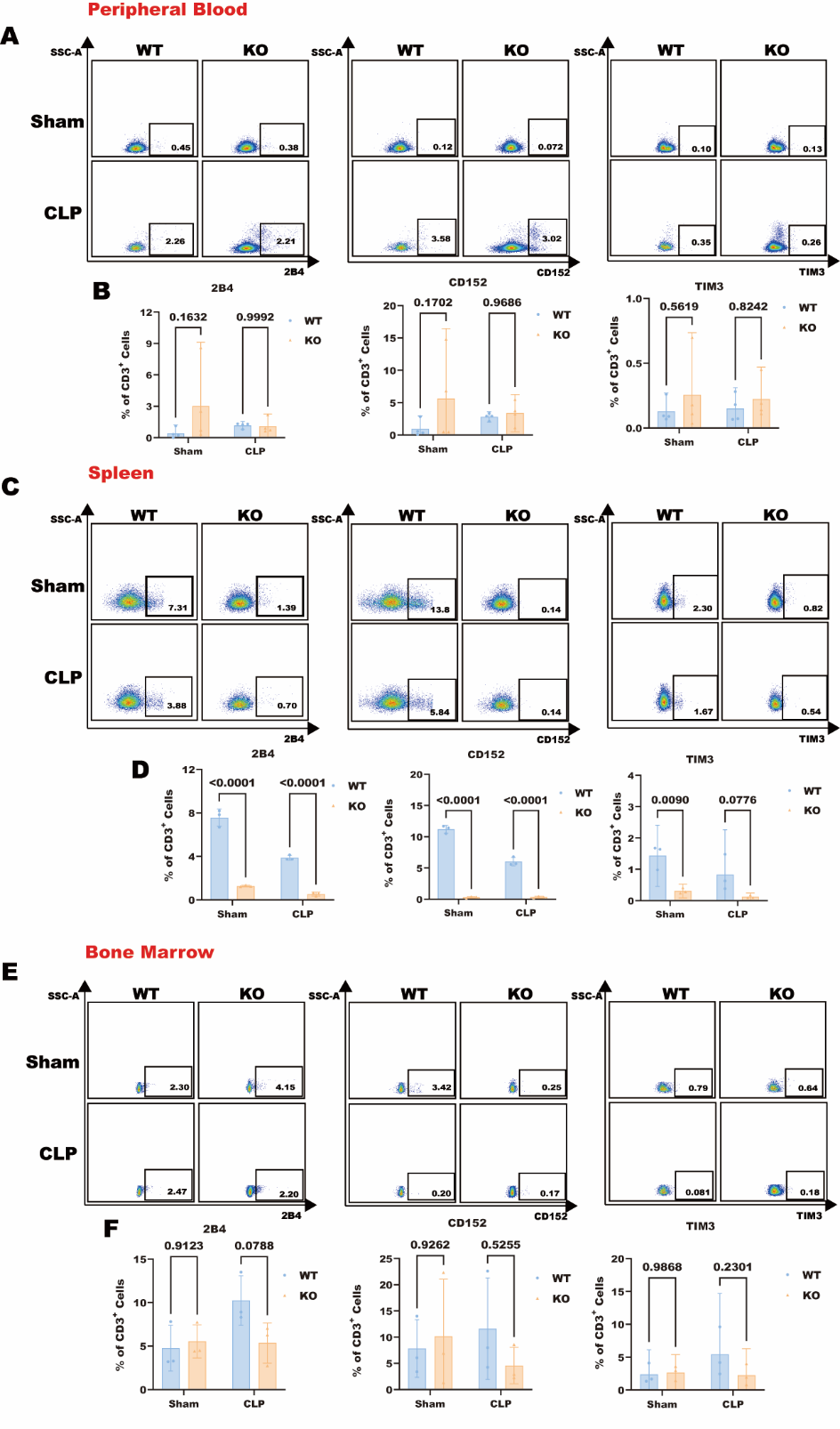

**Expression of 2B4, Cd152 and Tim3 in mice.** Flow analysis shows expression of 2B4, Cd152 and Tim3 in peripheral blood (A), spleen (C) and bone marrow (E) T cells of wild type and knockout mice. Statistics was performed using 2way ANOVA test in (B), (D) and (F). Data are shown as mean ± SD.

**Fig. S14**

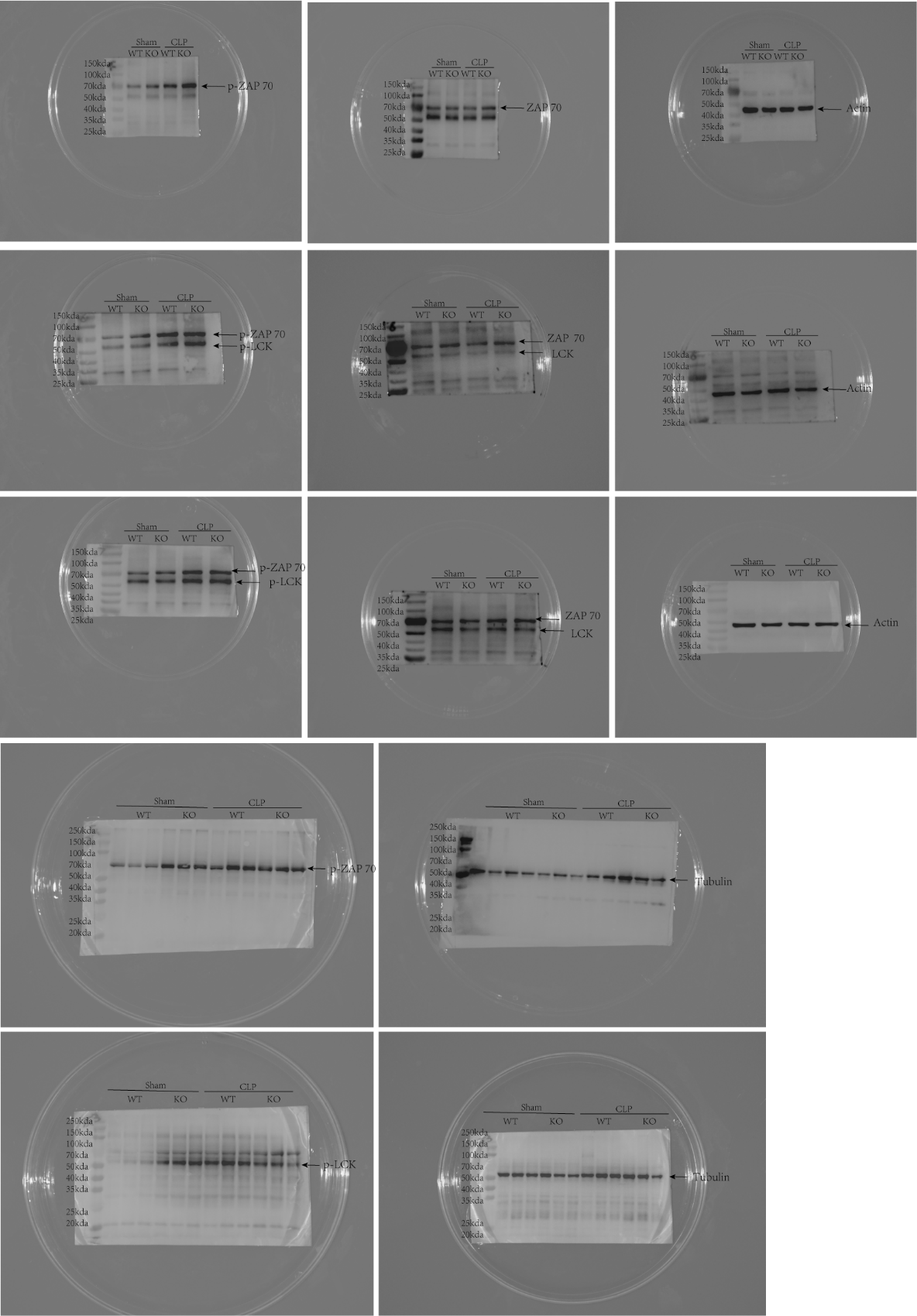

**Duplicates of western blot for phosphorylation of ZAP70 and LCK detection.**

**Fig. S15**

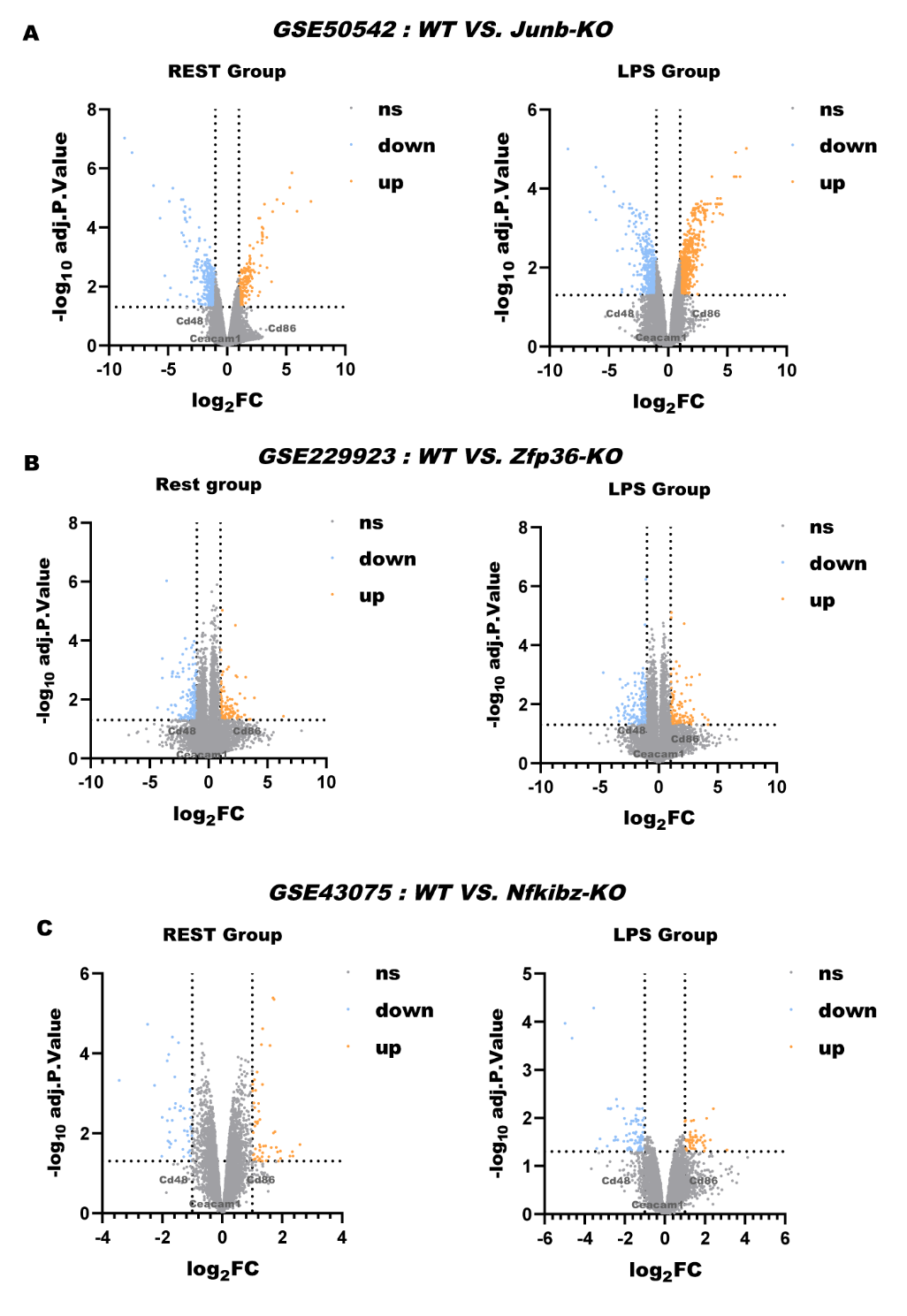

**Volcano plots for genes in REST and LPS group.** (A) GSE50542: WT VS. Junb-KO; (B) GSE229923: WT VS. Zfp36-KO; (C) GSE43075: WT VS. Nfkibz-KO.
